# Safety and immunogenicity of a reduced dose of the BNT162b2 mRNA COVID-19 vaccine (REDU-VAC): a single blind, randomized, non-inferiority trial

**DOI:** 10.1101/2022.03.25.22272599

**Authors:** Pieter Pannus, Stéphanie Depickère, Delphine Kemlin, Sarah Houben, Kristof Y Neven, Leo Heyndrickx, Johan Michiels, Elisabeth Willems, Stéphane De Craeye, Antoine Francotte, Félicie Chaumont, Véronique Olislagers, Alexandra Waegemans, Mathieu Verbrugghe, Marie-Noëlle Schmickler, Steven Van Gucht, Katelijne Dierick, Arnaud Marchant, Isabelle Desombere, Kevin K Ariën, Maria E Goossens

**Affiliations:** Scientific Direction Infectious Diseases in Humans, Sciensano, Brussels, Belgium; Institute for Medical Immunology and ULB Centre for Research in Immunology (U-CRI), Université libre de Bruxelles (ULB), Gosselies, Belgium; Virology Unit, Department of Biomedical Sciences, Institute of Tropical Medicine, Antwerp, Belgium; Mensura EDPB, Occupational Health Service, Antwerp, Belgium; Department of Biomedical Sciences, University of Antwerp, Antwerp, Belgium

**Keywords:** COVID-19, fractional dose, reduced dose, mRNA vaccination, non-inferiority

## Abstract

1

**Background:** The use of fractional dose regimens of COVID-19 vaccines has the potential to accelerate vaccination rates in low-income countries. Dose-finding studies of the mRNA vaccine BNT162b2 (Pfizer-BioNTech) have suggested that a fractional dose induces comparable antibody responses to the full, licensed dose in people below 55 years old. Here, we report the safety and immunogenicity of a fractional dose regimen of the BNT162b2 vaccine.

**Methods:** REDU-VAC is a participant-blinded, randomised, phase 4, multicentre, non-inferiority study investigating safety, reactogenicity and immunogenicity of BNT162b2. Adults aged between 18 and 55 years, without uncontrolled co-morbidities, either previously infected or infection naïve, were eligible and recruited at five sites across Belgium. Participants were randomly assigned to receive 20µg/20µg (fractional dose) or 30µg/30µg (full dose) of BNT162b2, administered intra-muscularly at a three-week interval. The primary endpoint was the geometric mean ratio (GMR) of serum SARS-CoV-2 anti-RBD IgG titres at 28 days post second dose between the reduced and the full dose regimens. The reduced dose was considered non-inferior to the full dose if the lower limit of the two-sided 95% CI of the GMR was greater than 0.67. The primary analysis was done on the per-protocol population, including infection naïve participants only.

**Findings:** Between April 19 and April 23, 2021, 145 participants were enrolled in the study and randomized, of whom 141 were vaccinated and reached the primary endpoint. Participants were mostly female (69.5%), of European origin (95%), with a mean age of 40.4 years (SD 7.9). At 28 days post second dose, the geometric mean titre (GMT) of SARS-CoV-2 anti-RBD IgG of the reduced dose regimen (1,705 BAU/mL) was not non-inferior to the full dose regimen (2,387 BAU/mL), with a GMR of 0.714 (two-sided 95% CI 0.540-0.944). No serious adverse events occurred.

**Conclusions:** While non-inferiority of the reduced dose regimen was not demonstrated, the SARS-CoV-2 anti-RBD IgG titre was only moderately lower than that of the full dose regimen and, importantly, still markedly higher than the reported antibody response to the licensed adenoviral vector vaccines. These data suggest that reduced doses of the BNT162b2 mRNA vaccine may offer additional benefit as compared to the vaccines currently in use in most low and middle-income countries, warranting larger immunogenicity and effectiveness trials. The trial is registered at ClinicalTrials.gov (NCT04852861).

## Introduction

Today, less than 15% of people in low-income countries (LIC) have been vaccinated against COVID-19 with at least one dose, compared to 68% in high-income countries (1). Besides allowing the emergence of new SARS-CoV-2 variants and their subsequent spread around the world, this blatant inequity in global vaccine distribution is leaving hundreds of millions of people vulnerable to the deadly virus. According to the UN, vaccine inequity will further deepen inequality and have a lasting impact on socio-economic recovery in LIC (2).

The lack of access to COVID-19 vaccines is partly due to a phenomenon called ‘vaccine nationalism’, wherein most vaccines are being reserved for wealthy countries, and vaccine-producing countries limit exports to make sure that their own population is vaccinated first. Another important factor is the cost. Under current pricing, LIC would have to increase health care expenditure by up to 50%, compared to less than 1% for high-income countries, to vaccinate 70% of their population (3–5). The WHO, Gavi and CEPI co-led program COVAX was set up to stimulate global equitable access to COVID-19 vaccines. Although this program has led to the donation of over 1 billion vaccine doses to LIC to date, more doses, better logistics, and political will are required to cover the needs of the Global South.

Fractionating doses as a dose-sparing strategy might help speed up vaccination rates by effectively reducing the cost per dose and thereby increasing the number of available doses. Based on proven safety and immunogenicity, the World Health Organization (WHO) has recommended fractional doses in the past during outbreaks of yellow fever, polio and meningococcal disease in case of vaccine shortages in resource-limited settings (6–9).

In terms of fractional vaccine doses for COVID-19 however, data is mostly limited to dose-finding studies. In a phase 1/2 trial in healthy adults aged 18-55 years, humoral and cellular responses to the BNT162b2 mRNA vaccine (Comirnaty®, Pfizer-BioNTech) were found to be very similar between the full dose of 30µg and fractional doses of 20µg and even 10µg (10). For the mRNA-1273 vaccine (Spikevax®, Moderna), a phase 2 trial showed comparable humoral responses (binding and neutralizing antibody titres) to a 50µg dose as compared to the full dose of 100µg (11). As far as we know, there has only been one trial reporting on clinical efficacy of fractional dosing in a small number of subjects. This multicentre trial observed 67%-97% efficacy of protection against symptomatic COVID-19 in a sub-group of participants who were primed with a half dose and boosted with a full dose of the ChAdOx1 nCoV-19 (Vaxzevria®, AstraZeneca) vaccine (12). In a recent paper, Więcek *et al.* used modelling data by Khoury *et al.* to predict vaccine efficacy of fractional doses (13, 14). Compared to the 95% efficacy provided by the full dose of 30µg, fractional doses of 10µg and 20µg of BNT162b2 are estimated to still be 80-90% effective in adults below 55 years old. Together with the dose-sparing advantage, the possibly fewer side effects caused by lower dosing, as suggested by some clinical data, may help tip the balance in favour of implementing fractional dosing strategies (15).

As more data is urgently needed to fill this knowledge gap and guide the potential use of fractional doses of mRNA vaccines, we conducted a randomised controlled trial to determine whether immune responses to a reduced dose of BNT162b2 (20µg) are non-inferior to the full, licensed dose (30µg). We found that binding and neutralizing antibody responses to the reduced dose of 20µg were indeed inferior to the full dose of 30µg of BNT162b2, but nevertheless still markedly higher than responses induced by other approved COVID-19 vaccines with proven efficacy.

## Methods

### Study design

REDU-VAC is a participant-blinded, randomised, phase 4, multicentre, non-inferiority study investigating safety, reactogenicity and immunogenicity of a fractional dose of the mRNA COVID-19 vaccine BNT162b2 (Pfizer-BioNTech). Recruitment occurred at five sites across Belgium of the ‘External Department of Prevention and Protection at work’ of Mensura. The trial was reviewed and approved by the Erasme Hospital Ethics Committee (P2021/251) and the Federal Agency for Medicines and Health Products (EudraCT: 2021-002088-23A). The trial was registered at ClinicalTrials.gov (NCT04852861). The protocol is available in S1 Appendix.

Two prime-boost schedules are compared consisting, respectively, of two doses of 20µg (reduced dose schedule) and two doses of 30µg (full dose schedule) of BNT162b2. All participants were randomly assigned to receive either the reduced or the full dose schedule.

### Participants

Non-previously vaccinated adults aged 18-55 years old who were in good general health or having well-controlled co-morbidities, either infection naïve or previously infected, were eligible. Exclusion criteria included history of severe adverse reactions associated with a vaccine or severe allergic reaction to any component of the study intervention, acute severe febrile illness or acute infection, pregnancy and breastfeeding.

### Randomisation and masking

Participants were block-randomised (random block size between 2, 4, 6, and 8, ratio of 1:1:1:1) within female/20µg, female/30µg, male/20µg, and male/30µg using the blockrand R package by the study statistician (16). The age distribution in each group was then verified to be not different in the four groups using a Kruskal-Wallis test. The obtained p-value was 0.54.

Participants and laboratory staff were masked to the administered vaccine dose (20µg versus 30µg). To ensure participant blinding to the vaccine dose, randomisation lists were kept out of sight, vaccines were prepared, and syringes were filled beforehand.

### Procedures

Participants were informed of the study and asked to register online before study participation. Those meeting all inclusion criteria were randomly assigned to one of the two study groups by the study statistician and were invited to the baseline visit (day 0) by the study nurses. At the baseline visit, participants provided informed consent before having blood drawn and being vaccinated.

Two different dosages of the same COVID-19 mRNA BNT162b2 vaccine were used in the study. BNT162b2 is a lipid nanoparticle–formulated, nucleoside-modified mRNA vaccine encoding a prefusion stabilized, membrane-anchored SARS-CoV-2 full-length spike protein. The 20µg and 30µg doses were administered via respectively a 0.2 mL and a 0.3 mL intra-muscular injection into the upper arm.

Vaccines were administered by trained study nurses at the different trial sites. Participants were observed for a minimum of 15 minutes after vaccination. The interval between the two vaccine doses was three weeks for all participants. Blood was drawn on days of vaccination (day 0 and day 21), four weeks post second dose (day 49), and six months post first dose (month six). One week after administration of each vaccine dose, participants were asked to record online any experienced local and systemic adverse events as well as their severity (mild/moderate/severe). Participants were also asked to record breakthrough infections by registering online the date of a positive SARS-CoV-2 molecular test (on the condition that no third vaccine dose had been received yet), and in case of symptomatic infection, the duration of symptoms. Breakthrough infections were followed-up until administration of a third vaccine dose.

SARS-CoV-2 anti-receptor binding domain (RBD) specific IgG concentrations were measured by ELISA (reported as Binding Antibody Units [BAU]/mL) on days 0/21/49 and month 6. Neutralizing antibody titres against SARS-CoV-2 Wuhan (2019-nCoV-Italy-INMI1, reference 169 008V-03893) (days 21/49), the B.1.617.2 Delta variant (83DJ-1) and the BA.1 Omicron variant (day 49) were measured with a live virus neutralization assay (VNA, reported as 50% neutralization, NT50) (17). The VNA was only performed against Delta and Omicron for samples with an NT50 (Wuhan) titre >50 and >400 respectively. Cellular responses were measured at day 49 both with IFN-γ enzyme-linked immunosorbent spot assay (ELISpot) as well as with multi-colour flow cytometry on a subsample of 45 randomly selected participants. Detailed methods are described in the S2 appendix.

Previous infection status was established following a decision tree (S1 Fig). Participants with a previous laboratory-confirmed SARS-CoV-2 infection were considered previously infected irrespective of baseline serology. All other participants with a baseline anti-RBD IgG titre <5 BAU/mL were considered infection naïve, and those ≥ 30 BAU/mL were considered previously infected. Participants with a titre ≥ 5 and < 30 BAU/mL were further tested with a different, multiplexed immunoassay (Multi-SARS-CoV-2 Immunoassay) detecting four targets: RBD, spike subunit 1 (S1), spike subunit 2 (S2) and nucleocapsid (N) (17). Participants with ≥3 out of 4 targets positive were considered previously infected.

### Outcomes

The primary outcome was serum SARS-CoV-2 anti-RBD IgG concentration 28 days post second dose (day 49) in infection naïve participants. Secondary outcomes included reactogenicity, as measured by reported local and systemic adverse events in the week following each vaccination, and safety, as measured by suspected unexpected serious adverse reactions, serious adverse reactions, and adverse reactions with grade equal or more than three over the entire study period.

Immunological secondary outcomes include SARS-CoV-2 anti-RBD IgG at day 0, day 21 and month 6; VNA titres at days 21 (Wuhan) and 49 (Wuhan, Delta and Omicron); and SARS-CoV-2 specific cell frequencies at day 49 in infection naïve participants only as well as in infection naïve and previously infected participants combined.

### Statistical analysis

The sample size was calculated assuming a true difference of geometric means of the primary outcome on the log_10_ scale being 0 between the reduced and the full dose, and a standard deviation of GMT on the log_10_ scale being 0.27 (17). A minimum of 50 naïve participants per arm was necessary to achieve 90% power at a two-sided 5% significance level. The geometric mean ratio (GMR) was then calculated as the anti-logarithm of the difference between the mean on the log_10_ scale of the primary outcome in the reduced dose and that in the full dose (reference dose), after adjusting by a mixed-effect linear model using age, gender, and baseline titre of SARS-CoV-2 anti-RBD IgG as fixed factors and study sites as random factor. To conclude on non-inferiority between the two groups, the WHO criterion of 0.67 was used (18), i.e. the reduced dose was considered non-inferior when the lower limit of the two-sided 95% CI of the GMR was greater than this cut-off. In the same way, GMRs were also calculated at day 49 for VNA titres against Wuhan and Delta variants.

The proportion of participants with responses higher than the lower limit of detection were calculated for SARS-CoV-2 anti-RBD IgG at days 0, 21 and 49, and for VNA titres at days 21 (Wuhan) and 49 (Wuhan, Delta, Omicron) with 95% CI calculated by the binomial exact method. They were compared between the reduced and the full dose groups using the Fisher’s exact test. Data below the lower limit of detection were given a value equal to half of the threshold before transformation. Comparisons of primary and secondary outcomes were evaluated by linear mixed-effect model adjusting for age, gender, and baseline titre of SARS-CoV-2 anti-RBD IgG (for days 21 and 49) or naïve/not naïve category (for day 0) as fixed factors and study sites as random factor. Correlations between SARS-CoV-2 anti-RBD IgG and VNA titres (Wuhan), and between VNA against Wuhan and Delta variants were evaluated by Pearson correlation coefficients. Concerning cellular responses, the proportion of participants with responses higher than 0 in IFN-γ ELISpot and with a positive response in flow cytometry was computed at day 49 with 95% CI calculated by the binomial exact method. Fisher’s exact test was used to compare reduced and full dose groups. For IFN-γ ELISpot analysis, data equal to 0 were given a value of 1 before transformation, and for flow cytometry, negative frequencies were given a value of 0.0001 before transformation. Comparisons of GMTs were evaluated by linear mixed-effect model adjusting for age, gender, and baseline titre of SARS-CoV-2 anti-RBD IgG as fixed factors and study sites as random factor. All statistical analyses were done using R version 4.1.2.

## Results

Between April 19 and April 23, 2021, 152 participants were screened for eligibility in five sites across Belgium among whom 145 were enrolled in the study and randomized (Fig 1). Four participants were excluded from the study due to pregnancy, active infection, high-risk contact or medical reasons. Participants were mostly female (69.5%), of European origin (95%), with a mean age of 40.4 years (SD 7.9). Six participants (4.3%) were taking medication to treat a co-morbidity. Baseline characteristics were well balanced across the two study arms (Table 1). Hundred twenty-four participants were considered SARS-CoV-2 infection naïve and 17 were previously infected at baseline. Here, we focus on the results of the per-protocol (PP) analysis including infection naïve participants only and present the results of the intention-to-treat (ITT) analysis, including both naïve and previously infected participants, in supplementary data.

**Fig 1.**
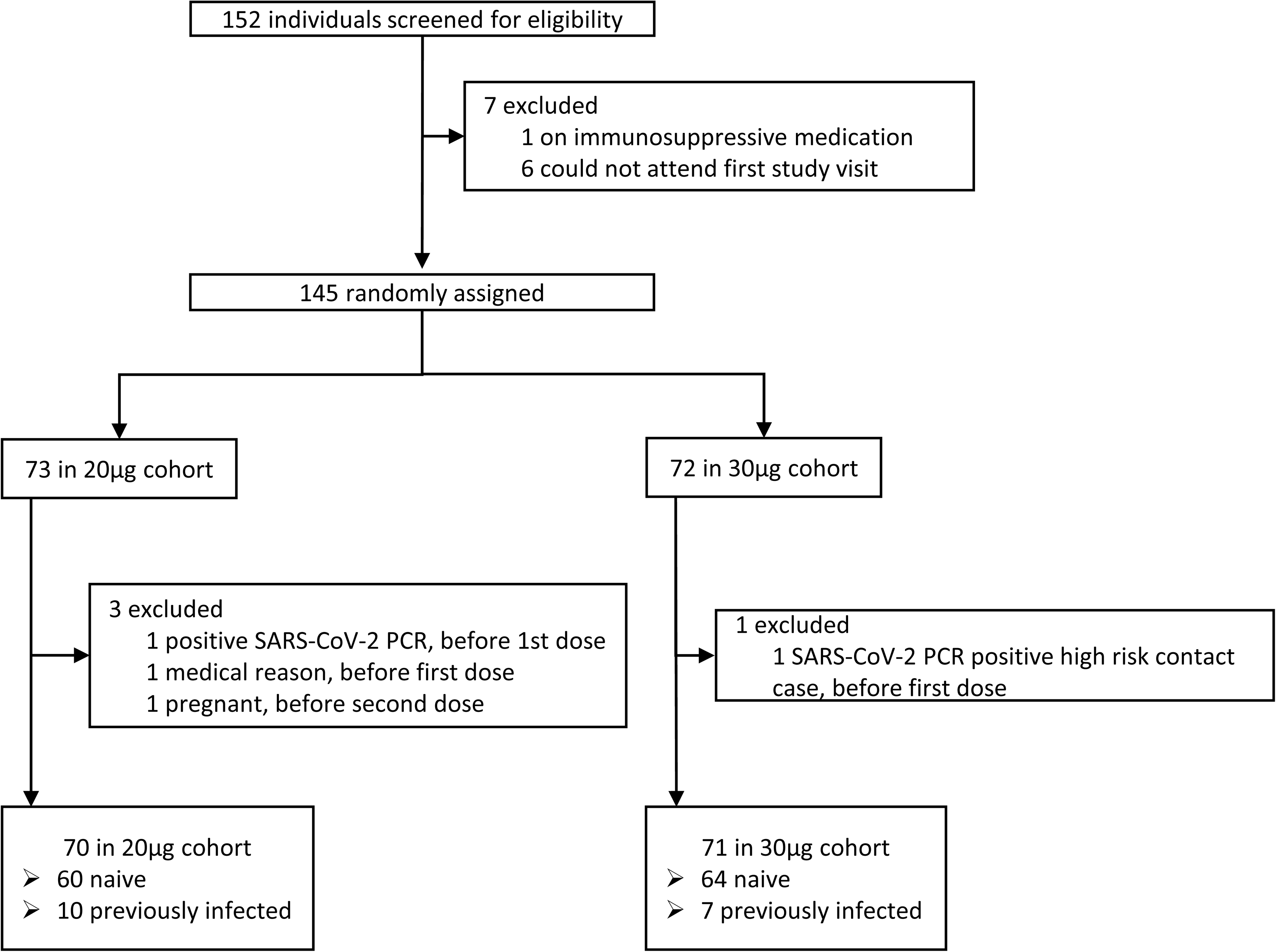
Trial profile.

**Table 1.** Baseline characteristics by cohort and study arm.

|  | Per-protocol |  | Intention-to-treat |  |
| --- | --- | --- | --- | --- |
|  | 20 µg | 30 µg | 20 µg | 30 µg |
| Participants, n | 60 | 64 | 70 | 71 |
| Age, years |  |  |  |  |
| Mean (SD) | 40.4 (7.5) | 41.0 (8.2) | 39.8 (7.9) | 41.0 (8.0) |
| Median (range) | 39.5 (23.0-54.0) | 41.5 (24.0-55.0) | 39.0 (23.0-54.0) | 41.0 (24.0-55.0) |
| Sex |  |  |  |  |
| Female | 43 (71.7%) | 43 (67.2%) | 49 (70.0%) | 49 (69.0%) |
| Male | 17 (28.3%) | 21 (32.8%) | 21 (30.0%) | 22 (31.0%) |
| Ethnicity |  |  |  |  |
| European | 56 (93.3%) | 63 (98.4%) | 65 (92.9%) | 69 (97.2%) |
| Latin | 1 (1.7%) | 0 | 1 (1.4%) | 0 |
| Asian | 1 (1.7%) | 0 | 1 (1.4%) | 1 (1.4%) |
| Sub-Saharan Africa | 2 (3.3%) | 0 | 2 (2.9%) | 0 |
| Northern-Africa | 0 | 1 (1.6%) | 1 (1.4%) | 1 (1.4%) |
| Comorbidities |  |  |  |  |
| Cardiovascular | 1 (1.7%) | 2 (3.1%) | 2 (2.9%) | 2 (2.8%) |
| Oncological | 0 | 1 (1.6%) | 0 | 1 (1.4%) |
| Respiratory | 0 | 1 (1.6%) | 0 | 1 (1.4%) |
| BMI |  |  |  |  |
| <18.5 | 1 (1.7%) | 1 (1.6%) | 1 (1.4%) | 1 (1.4%) |
| ≥18.5 and <25 | 39 (65.0%) | 36 (56.3%) | 45 (64.3%) | 40 (56.3%) |
| ≥25 | 20 (33.3%) | 27 (42.1%) | 24 (34.3%) | 30 (42.3%) |

Data are n (%) unless otherwise indicated.

At the primary endpoint, 28 days post second dose, the GMT of SARS-CoV-2 anti-RBD IgG in the PP cohort was 1705 BAU/ml in the 20µg group and 2387 BAU/ml in the 30µg group (Table 2). The GMR of 0.714 (0.540-0.944, 95% CI) indicated that non-inferiority was not demonstrated given that the lower limit of the 95% CI was inferior to the WHO recommended margin of 0.67. Similarly, the ITT analysis did not demonstrate non-inferiority of the reduced dose either, with GMTs of 1822 BAU/ml and 2381 BAU/ml in the 20µg and 30µg arms, respectively, and a GMR of 0.765 (0.593-0.987, 95% CI) (S1 Table).

**Table 2.**
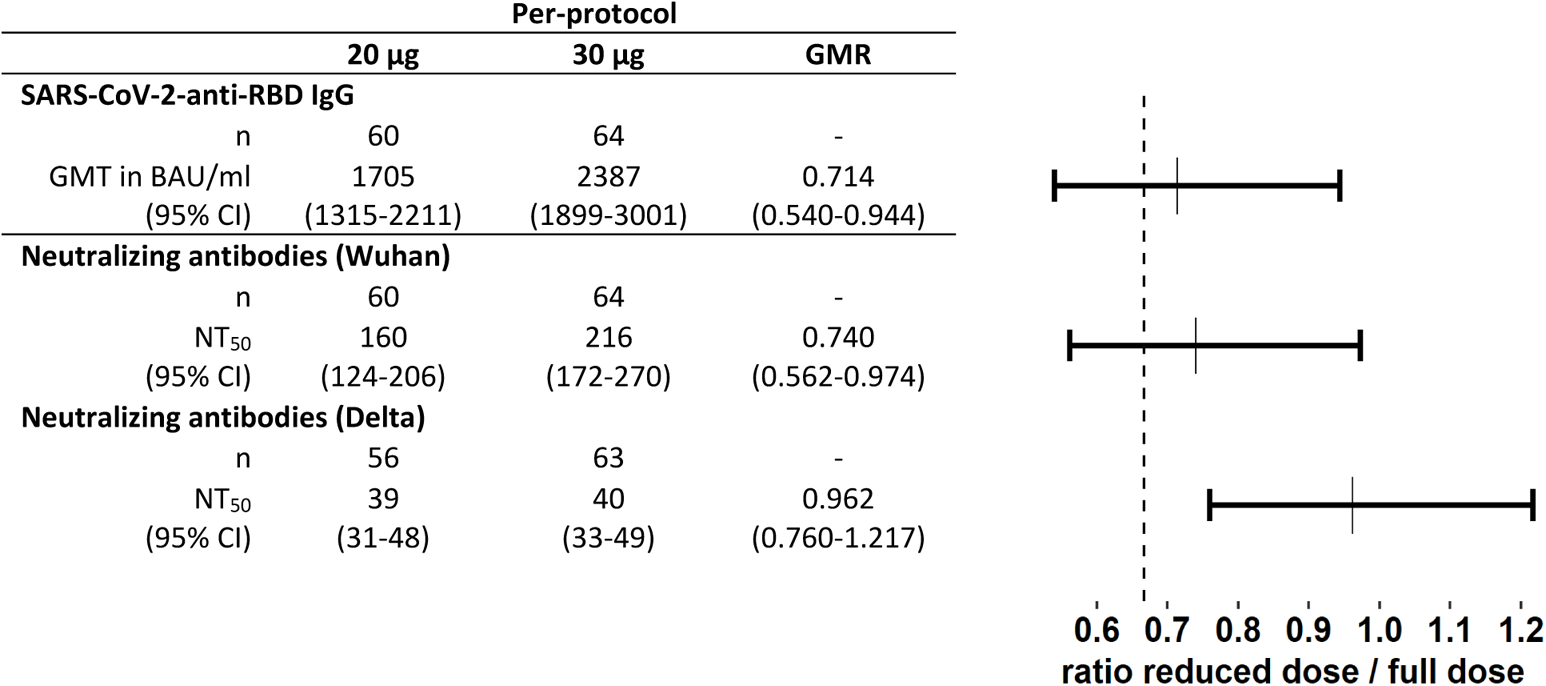
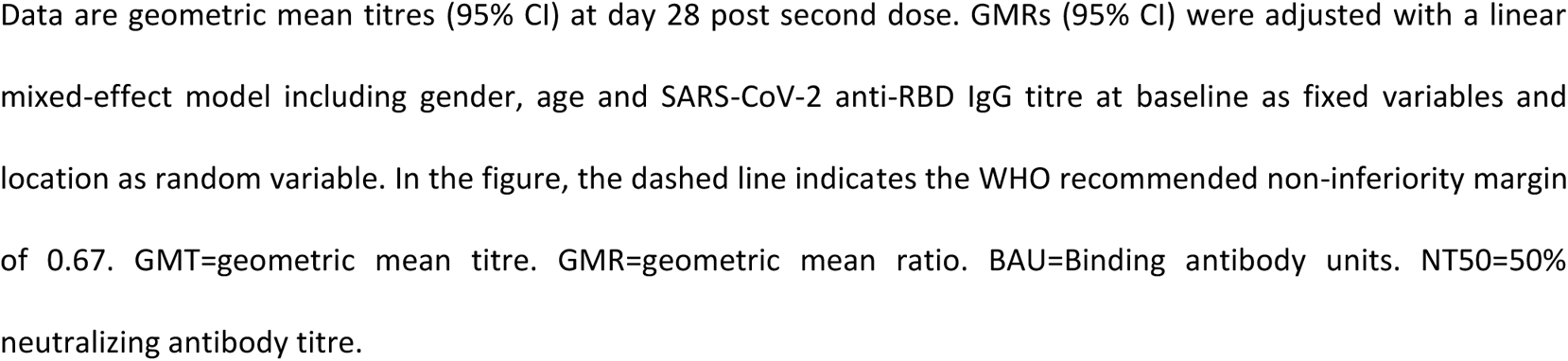
Immune responses by study arm at 28 days post second vaccine dose (Day 49) and non-inferiority analysis in the per-protocol cohort.

We observed strong correlations between SARS-CoV-2 binding (anti-RBD IgG) and neutralizing (Wuhan NT_50_) antibody titres in both the reduced and the full dose arms (Fig 2A). As a result, GMRs for neutralizing titres were similar to those described above for binding titres. Indeed, GMTs of neutralizing antibody titres against Wuhan were 160 (124-206) and 216 (172-270) in the 20µg and 30µg arms, respectively, with a GMR of 0.740 (0.562-0.974) (Table 2). In the ITT analysis, GMTs were 216 (95% CI: 170-276) and 279 (224-347) in the 20µg and 30µg arms, respectively, with a GMR of 0.776 (0.592-1.017) (S1 Table). In both analyses, non-inferiority was not demonstrated.

**Fig 2.**
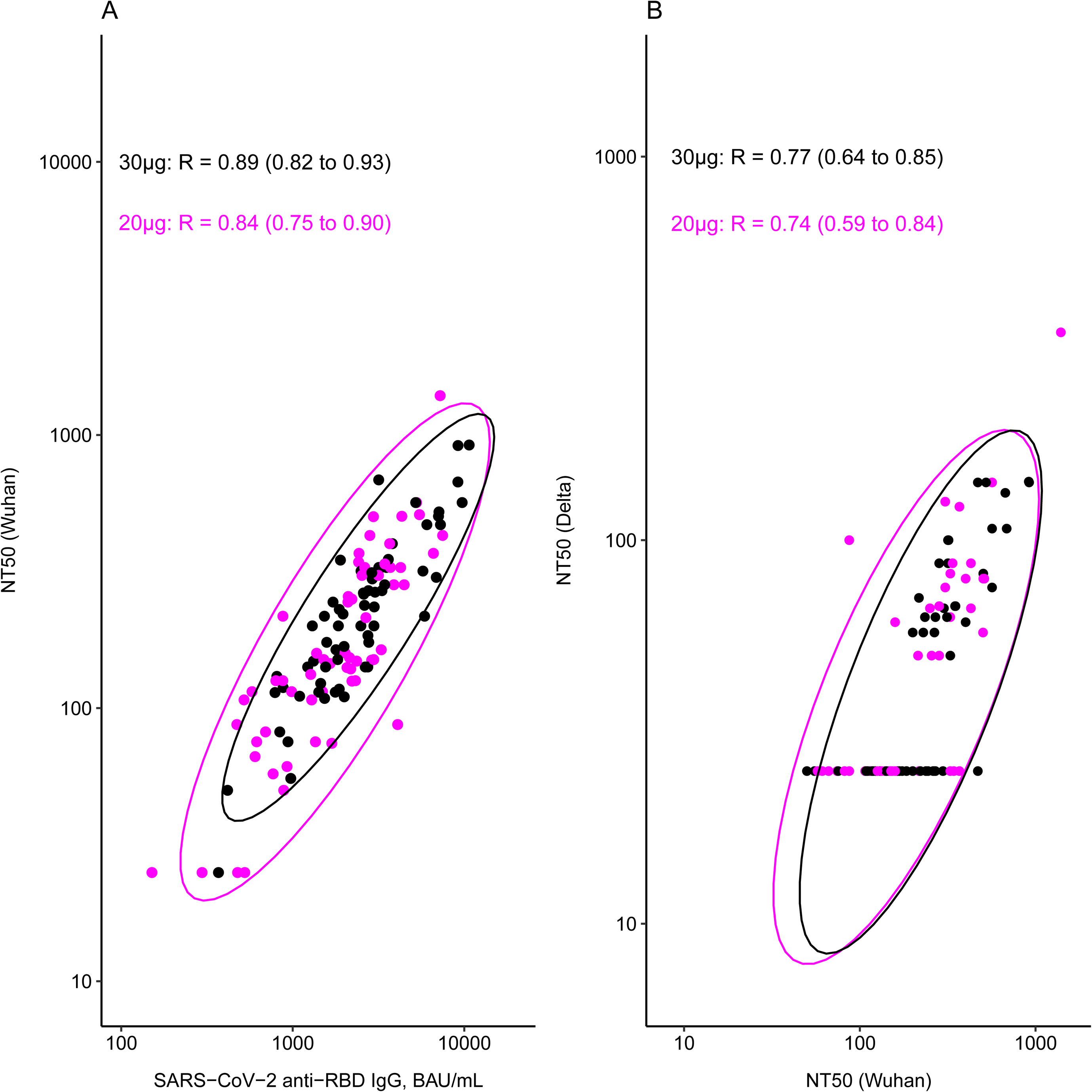
Correlations between immune responses per study arm. Correlations were analysed at 28 days after the second vaccine dose between SARS-CoV-2 anti-RBD IgG binding antibodies and SARS-CoV-2 Wuhan neutralizing antibodies (A), and between SARS-CoV-2 Wuhan and Delta variant neutralizing antibodies (B). Pearson correlation coefficients (95% CI) are given per study arm. Ellipses represent the 95% CI for the two study arms (purple=20µg, black=30µg), assuming multivariate normal distributions. NT50=50% neutralizing antibody titre, RBD=SARS-CoV-2 receptor binding domain, BAU=binding antibody units.

Neutralizing antibodies against Delta and Omicron were only tested for participants with an NT50 (Wuhan) >50 and >400, respectively. As expected, neutralizing capacity against these variants was much lower compared to Wuhan. In the samples tested, just 21/56 (38%) and 25/63 (40%) naïve participants had detectable Delta neutralizing titres in the 20µg and 30µg arms, respectively (Table 3). For Omicron, none of the naïve participants (0/8 and 0/11 for 20µg and 30µg, respectively) had detectable neutralizing activity. Titres against Wuhan and Delta were strongly correlated in both study arms (Fig 2B). Correlation between Wuhan and Omicron could not be determined due to the mostly undetectable titres. Non-inferiority of the reduced dose to the full dose was demonstrated for Delta neutralizing titres, in the PP and ITT analyses, as the lower limit of the 95% CIs were superior to the WHO margin of 0.67 (Table 2 and S1 Table). As all Omicron neutralizing titres were undetectable in naïve participants, regardless of the study arm, a non-inferiority analysis was not performed.

**Table 3.** Humoral and cellular responses by study arm in the per-protocol cohort at the different time points.

| Per-protocol |  |  |  |  |
| --- | --- | --- | --- | --- |
|  |  | 20 µg | 30 µg | p-value |
| **SARS-CoV-2 anti-RBD IgG** |  |  |  |  |
| **Day 0** |  |  |  |  |
|  | n | 60 | 64 |  |
| Concentration (BAU/ml) |  | 2.6<br>(2.3-2.9) | 2.8<br>(2.5-3.0) | p=0.421 |
| > 5 BAU/ml |  | 2<br>(3%, 0.4-12) | 4<br>(6%, 1.7-15) | p=0.681 |
| **Day 21** |  |  |  |  |
|  | n | 60 | 64 |  |
| Concentration (BAU/ml) |  | 117<br>(84-162) | 223<br>(166-298) | p<0.001 |
| > 5 BAU/ml |  | 59<br>(98%, 91-100) | 64<br>(100%, 94-100) | p=0.484 |
| **Day 49** |  |  |  |  |
|  | n | 60 | 64 |  |
| Concentration (BAU/ml) |  | 1705<br>(1315-2211) | 2387<br>(1899-3001) | p=0.019 |
| > 5 BAU/ml |  | 60<br>(100%, 94-100) | 64<br>(100%, 94-100) | p=1.000 |
| **Month 6** |  |  |  |  |
|  | n | 50 | 60 | p=0.007 |
| Concentration (BAU/ml) |  | 157<br>(113-217) | 246<br>(188-323) |  |
| > 5 BAU/ml |  | 50<br>(100%, 93-100) | 60<br>(100%, 94-100) | p=1.000 |
| **Neutralizing antibodies** |  |  |  |  |
| **Wuhan - Day 21** |  |  |  |  |
|  | n | 60 | 64 |  |
| NT<sub>50</sub> |  | 27<br>(24-31) | 29<br>(26-33) | p=0.355 |
| > 50 |  | 7<br>(12%, 5-23) | 10<br>(16%, 8-27) | p=0.606 |
| **Wuhan - Day 49** |  |  |  |  |
|  | n | 60 | 64 | - |
| NT<sub>50</sub> |  | 160<br>(124-206) | 216<br>(172-270) | p=0.032 |
| > 50 |  | 56<br>(93%, 84-98) | 63<br>(98%, 92-100) | p=1.000 |
| **Delta - Day 49** |  |  |  |  |
|  | n | 56 | 63 |  |
| NT<sub>50</sub> |  | 39<br>(31-48) | 40<br>(33-49) | p=0.743 |
| > 50 |  | 21 | 25 | p=0.852 |

|  |  | (38%, 25-51) | (40%, 28-53) |  |
| --- | --- | --- | --- | --- |
| **Omicron - Day 49** |  |  |  |  |
|  | n | 8 | 11 |  |
|  | NT<sub>50</sub> | 25<br>(25-25) | 25<br>(25-25) | - |
|  | > 50 | 0<br>(0%, 0-37) | 0<br>(0%, 0-28) | p=1.000 |
| **IFN-γ producing cells per million PBMCs** |  |  |  |  |
| **SARS-CoV-2 S1-specific - Day 49** |  |  |  |  |
|  | n | 18 | 24 |  |
|  | Mean number of cells/million | 71.5<br>(33.0-155.1) | 105.9<br>(60.0-187.0) | p=0.327 |
|  | > LOD | 12<br>(67%, 41-87) | 19<br>(79%, 58-93) | p=0.812 |
| **SARS-CoV-2 S2-specific - Day 49** |  |  |  |  |
|  | n | 18 | 24 |  |
|  | Mean number of cells/million | 88.5<br>(34.0-230.3) | 118.3<br>(50.9-274.8) | p=0.509 |
|  | > LOD | 11<br>(61%, 36-83) | 16<br>(67%, 45-84) | p=1.000 |

Data are geometric means (95% CI) for continuous variables, and n (%; 95% CI) for binary values. The LLOQ for the SARS-CoV-2 anti-RBD IgG was 5 BAU/mL, NT<sub>50</sub> = 50 for the live virus neutralisation assay, and 54 and 66 cells/million PBMCs for S1 and S2, respectively, for ELISpot. For continuous variables, p-values are reported using a linear mixed-effect model adjusted for gender, age and baseline infection status (for day 0 data) or baseline SARS-CoV-2 anti-RBD IgG titre (for day 21/49 and month 6 data) as fixed variables and location as random variable. Fisher's exact test was used to report p-values for binary variables. BAU=Binding antibody units. NT<sub>50</sub>=50% neutralizing antibody titre. PBMC=peripheral blood mononuclear cell. S1=SARS-CoV-2 spike protein subunit 1. S2=SARS-CoV-2 spike protein subunit 2.

In both the reduced and full dose study arms, all participants had seroconverted by 28 days post second dose (Table 3, Fig 3A). However, the GMTs of anti-RBD IgG were significantly lower in the 20µg arm compared to the 30µg arm at the time of the second dose administration (day 21) and 28 days later (day 49). At six months post first dose anti-RBD IgG titres had waned significantly, but all participants remained seropositive. The GMT of anti-RBD IgG was still significantly lower in the reduced dose arm at this time point (Table 3, Fig 3A). In terms of neutralizing activity, 56/60 (93%) versus 63/64 (98%) of naïve participants from the 20µg arm and the 30µg arm, respectively, had detectable neutralizing antibody titres against Wuhan, with a significantly lower titre in the reduced dose versus full dose arm 28 days post second dose (Table 3, Fig 3B). Neutralizing antibodies against Delta and Omicron were only measured at day 49 and did not differ significantly between the two study arms (Table 3, Fig 3C/D).

**Fig 3.**
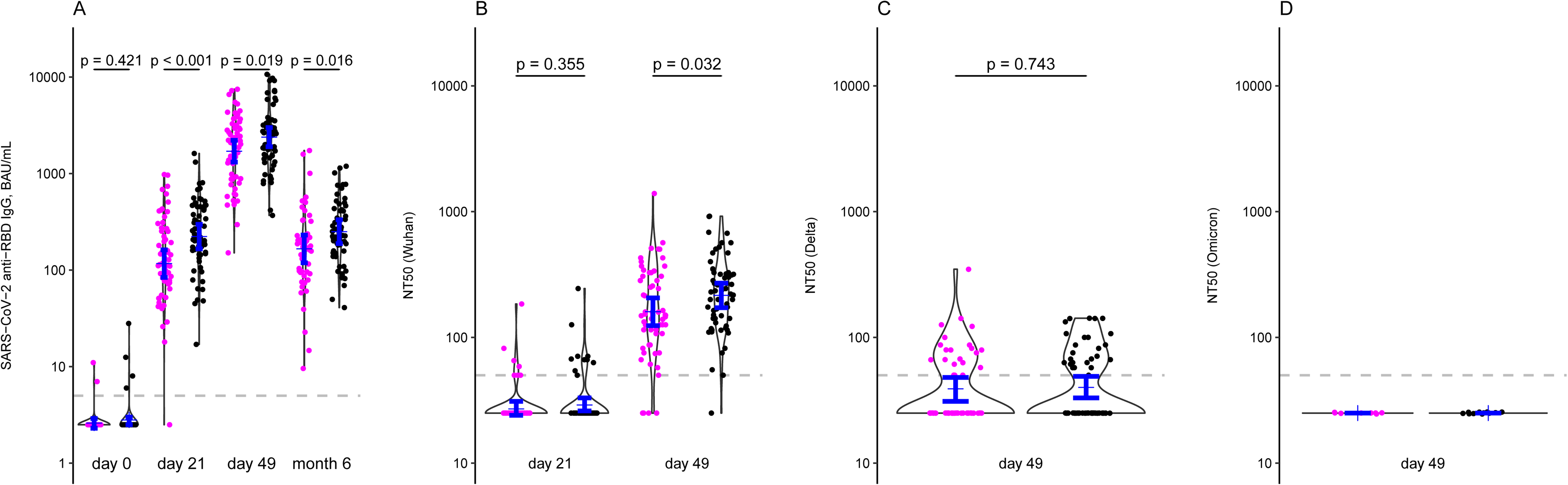
Kinetics of immune responses per study arm of the per-protocol cohort. SARS-CoV-2 anti-RBD IgG binding antibody data are shown on days 0/21/49 and month 6 post first dose (A), SARS-CoV-2 Wuhan neutralizing antibodies on days 21/49 (B) and SARS-CoV-2 Delta (C) and Omicron (D) neutralizing antibodies on day 49 per study arm (purple=20µg, black=30µg). In the case of SARS-CoV-2 anti-RBD IgG binding antibody at month 6, five samples were excluded due to a detected breakthrough infection before month 6. Blue bars indicate geometric mean titres with 95% CI. NT50=50% neutralizing antibody titre, RBD=SARS-CoV-2 receptor binding domain, BAU=binding antibody units.

Cellular responses elicited against a pool of SARS-CoV-2 spike protein subunit 1 (S1) and 2 (S2) peptides, measured with IFN-γ ELISpot, were not significantly different either between the reduced and full dose arms, either in the PP or the ITT cohort (Table 3, S2). The proportion of participants with detectable responses was also not significantly different, with 67% (41-87) versus 79% (58-93) for S1 in the 20µg and 30 µg arms, respectively, and 61% (36-83) versus 67% (45-84) for S2 in the 20µg and 30 µg arms, respectively (Table 3), in the PP cohort. Similar findings were observed in the ITT cohort (S2 Table). These data were corroborated by flow cytometry, where no differences were observed between the reduced and full dose arms in the proportion of S1 or S2 specific CD4+ and CD8+ T cells expressing CD154 (CD4+ T cells only), IFN-γ, IL-2, and TNF-α (S2 Fig).

A total of 18 breakthrough infections (BTI) were reported and all occurred between five to eight months after the first dose, coinciding with a major infection wave in the country due to the Delta variant (S3 Table). These BTIs occurred exclusively in infection naïve participants and were as likely to occur in the 20µg arm as in the 30µg arm (Fisher’s exact test, p=0.80). GMTs of binding and neutralizing antibodies at 28 days post second dose or six months post first dose were not significantly different between those experiencing and not experiencing a BTI (Wilcoxon rank sum test, p>0.14).

No serious adverse events were reported during the study period. No difference in frequency or severity of adverse events reported after each vaccine dose was observed between the 20µg and the 30µg arm (S4 Table, S3 Fig). The only exception was the reported severity of nausea after the second dose which was mostly moderate in the 20µg arm and mild in the 30µg arm.

## Discussion

The study results show that the administration of a reduced dose (2×20µg) of the mRNA vaccine BNT162b2 induces lower titres of SARS-CoV-2 anti-RBD IgG binding and SARS-CoV-2 Wuhan neutralizing antibodies than the administration of the full vaccine dose (2×30µg). No such difference was observed for SARS-CoV-2 Delta neutralizing antibodies but this can likely be explained by the overall lower titres as compared to Wuhan, with many participants having undetectable neutralizing antibody titres post vaccination. For Omicron neutralization, none of the naïve participants had detectable titres, precluding a proper comparison between study arms but highlighting the lack of induction of neutralizing capacity against the Omicron variant with two doses of BNT162b2.

The ratios of the GMTs across all analyses indicate that the investigated reduced dose regimen induced around 25-30% lower antibody titres than the licensed full dose regimen. The magnitude of this reduction and its potential impact on vaccine efficacy needs to be put into perspective, however. A recent non-inferiority trial compared immunogenicity of Pfizer-BioNTech’s BNT162b2 with Astra-Zeneca’s adenoviral ChadOx1 nCoV-19 vaccine, amongst other vaccination regimens. At 28 days post second dose, the GMT of anti-spike binding IgGs was 90% lower for ChadOx1 nCoV-19 than for BNT162b2 (19). Despite ChadOx1 nCoV-19’s markedly lower humoral responses, it still has been shown to have an efficacy against symptomatic infection of about 70%, and it has been approved for use in 182 countries with more than two billion doses administered, primarily in LIC (12, 20). We therefore argue that the moderately lower humoral response of the reduced dose regimen investigated in our study is still excellent, and likely provides significant protection against COVID-19 with additional benefit as compared to the non-mRNA vaccines currently in use in most low and middle-income countries (13, 14). Larger immunogenicity and effectiveness trials are warranted to support this notion.

In contrast to the humoral responses, we did not observe any significant differences in the cellular immune responses to vaccination. We used both ELISpot and flow cytometry to quantify SARS-CoV-2 spike protein subunits 1 and 2 specific cells, both showing very similar frequencies between study arms. This was not entirely unexpected, considering that differences in cellular responses have been reported before to be less pronounced than those in humoral responses (19). The equivalent cellular immune responses observed in this study further support the notion that this reduced dosage likely induces potent immunity.

While acknowledging that this trial was not designed nor powered to study efficacy of protection, we did not observe an obvious difference in incidence of breakthrough infections between both study arms. It is important to consider, however, that these infections were most likely caused by the Delta variant of SARS-CoV-2, around six months after vaccination, while neutralizing titres against the Delta variant at 28 days post second dose were already very low for both the reduced and full dose arms. In other words, a possible difference in protection may have gone unnoticed due to a generally low incidence of infection during the first months following vaccination.

Based on waning antibody data and the emergence of new variants, high-income countries are providing third doses to the general population, and several countries have started administering fourth doses to specific groups with comorbidities. In the meantime, primary vaccination coverage in LIC remains very low. The COVAX program was founded in April 2020 with the specific aim to provide global equitable access to COVID-19 vaccines. The program has delivered approximately one billion vaccine doses to 144 participating countries, but current production capacity for COVID-19 vaccines does not cover the global needs, thus delaying the end of the pandemic. We acknowledge that the use of fractional doses is associated with a number of programmatic and operational challenges which may hamper its roll-out, such as the possible need for special syringes, adjustments of diluent volumes, or exacerbated vaccine hesitancy due to lower antibody levels (22, 23). Nevertheless, we believe that these challenges are rather limited and can still be solved relatively quickly for the most part. This is in contrast to more structural strategies to speed up vaccination rates, such as strengthening logistics and building local vaccine production capacity, which are important but will arguably take more time to have an impact.

This study has several limitations, the first being the relatively limited sample size. A larger number of participants would have resulted in smaller confidence intervals around the GMRs, which might have impacted the conclusions on non-inferiority. Although males were underrepresented in this study, we do not believe this is a major limitation as immune responses to COVID-19 mRNA vaccination in healthy, younger subjects are only minimally gender-dependent and importantly, there is no basis to assume that fractional dosing would affect immune responses differently between males and females (24–26). Secondly, the small proportion of previously infected participants in our study population (17/144, 12%) does not allow for a separate sensitivity analysis in this group. With record high COVID-19 incidences worldwide, the proportion of the population who experienced a past infection is rapidly growing, making analyses including previously infected people ever more relevant. In addition, breakthrough infections were not actively monitored by regular molecular testing. Therefore, we may have missed asymptomatic infections, which are not reported by the study participants. Thirdly, while protection from infection or disease has been convincingly correlated with titres of binding and neutralizing antibodies, as discussed previously, it is not possible to determine with certainty that the moderately lower titres observed in our study will translate to equally moderately lower efficacy of protection (14, 27). Fourth, while the interval of three weeks between both vaccine doses is recommended by the manufacturer, larger intervals are commonly adopted and have been shown to induce higher immune responses as compared to the shorter three week interval (28, 29). Finally, a strength of the study in terms of global health relevance, is the relatively young age distribution of our cohort, ranging from 23 to 55 years old (median: 40). This makes our data especially relevant for the context where it can be applied most, i.e. LIC where age distribution is typically much lower than in high income countries (97% versus 81% of population below 65 years old, respectively) (30). Nevertheless, further studies are needed including participants who are more reflective of the target population in terms of genetic background and environment.

To our knowledge, this is the only non-inferiority study reporting on fractional dosing of a COVID-19 mRNA vaccine so far. Our study shows moderately lower humoral responses and similar cellular immune responses after two reduced doses of 20µg compared to two full doses of 30µg of the BNT162b2 mRNA vaccine. Nevertheless, antibody responses to the reduced dose remain far superior to what has been published in the literature so far for marketed adenoviral vector vaccines with proven efficacy of protection against disease. Considering this and the important potential in accelerating vaccination rates in LIC, our findings support the need for larger non-inferiority trials on fractional dosing for primary vaccination as well as for follow-up booster vaccinations. The relevance of our findings goes beyond the current pandemic, as they highlight the potential of fractional dosing as a dose-sparing strategy in future epidemics and vaccine shortages.

## Author Contributions

**Conceptualization:** Maria E Goossens, Isabelle Desombere, Kevin K Ariën, Arnaud Marchant, Pieter Pannus.

**Data Curation:** Kristof Y Neven, Stéphanie Depickère, Pieter Pannus, Sarah Houben.

**Formal Analysis:** Stéphanie Depickère, Pieter Pannus, Kristof Y Neven.

**Funding Acquisition:** Maria E Goossens, Katelijne Dierick

**Investigation:** Pieter Pannus, Kristof Y Neven, Leo Heyndrickx, Johan Michiels, Elisabeth Willems, Stéphane De Craeye, Antoine Francotte, Félicie Chaumont, Véronique Olislaghers, Delphine Kemlin, Alexandra Waegemans, Kevin K Ariën, Isabelle Desombere, Arnaud Marchant.

**Methodology:** Pieter Pannus, Maria E Goossens, Kevin K Ariën, Isabelle Desombere, Arnaud Marchant.

**Project Administration:** Maria E Goossens, Pieter Pannus.

**Resources:** Kristof Y Neven, Pieter Pannus, Mathieu Verbrugge, Marie-Noëlle Schmickler, Maria E Goossens, Kevin K Ariën, Isabelle Desombere, Arnaud Marchant.

**Software:** Stéphanie Depickère.

**Supervision:** Maria E Goossens, Pieter Pannus.

**Validation:** Pieter Pannus, Stéphanie Depickère, Kristof Y Neven, Kevin K Ariën, Isabelle Desombere, Arnaud Marchant.

**Visualization:** Stéphanie Depickère, Pieter Pannus.

**Writing – Original Draft Preparation**: Pieter Pannus, Stéphanie Depickère.

**Writing – Review & Editing:** Pieter Pannus, Stéphanie Depickère, Delphine Kemlin, Sarah Houben, Kristof Y Neven, Leo Heyndrickx, Véronique Olislagers, Alexandra Waegemans, Mathieu Verbrugghe, Marie-Noëlle Schmickler, Steven Van Gucht, Katelijne Dierick, Arnaud Marchant, Maria E Goossens, Isabelle Desombere, Kevin K Ariën.

## Data Availability

All data produced in the present study are available upon reasonable request to the authors.

## Acknowledgements

This study is funded by the Belgian Federal Government through Sciensano [COVID-19_SC004, COVID-19_SC059, COVID-19_SC095, COVID-19_SC096] and the Research Foundation - Flanders [grant number FWO G0G4220N to KKA]. DK is a Clinical Master Specialist Applicant to a Ph.D. of the Fonds de la Recherche Scientifique – FNRS. A.M. is Research Director of the FRS-FNRS, Belgium. We thank Martine Delaere, Kristine Massez, Isabelle Maufort and Jody Serré for their dedicated work as study nurses. We also thank Caroline Rodeghiero, Fabienne Jurion, Elfriede Heerwegh, Celien Van Oostveldt, Vincent Martens, Valérie Acolty, Inès Vu Duc, Sara Cuman, Robin Van Naemen, Maria Lara Escandell, Sandra Coppens, Ann Ceulemans and Koen Bartholomeeusen for their help in the laboratory, as well as Murat Gonen and Jonathan Masala for their logistical support. We thank Piet Maes (Rega Institute, KU Leuven, Belgium) to kindly provide the B.1.617.2 Delta variant (83DJ-1) isolate. Finally, we thank all study participants for their availability, flexibility and dedication to the study.

## SUPPLEMENTARY MATERIAL

**S1 Fig.**
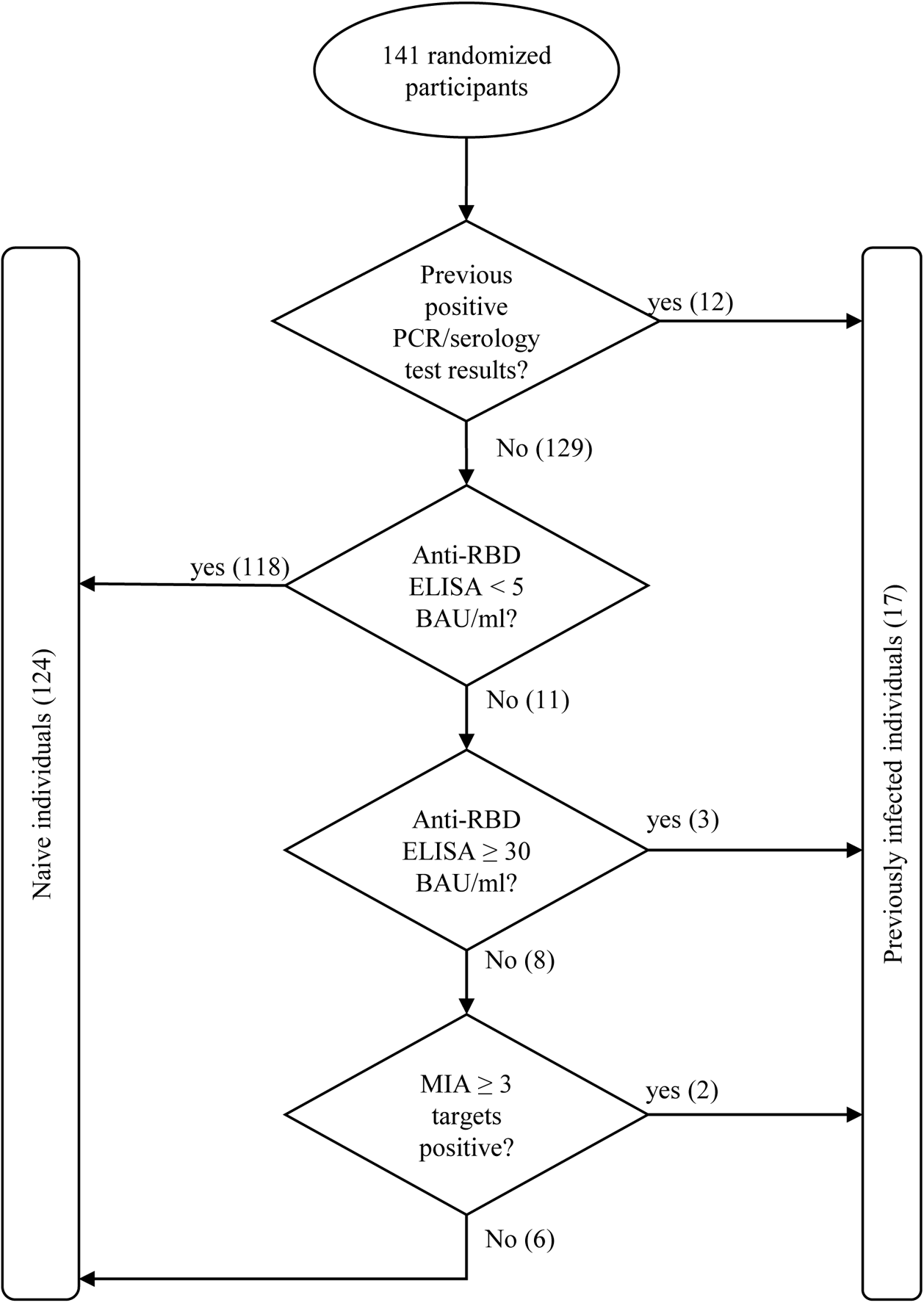
Decision tree to determine previous SARS-CoV-2 infection status at baseline. RBD=SARS-CoV-2 receptor binding domain, ELISA=Enzyme linked immunosorbent assay, BAU=binding antibody units, MIA=multiplex immunoassay.

**S1 Table.**
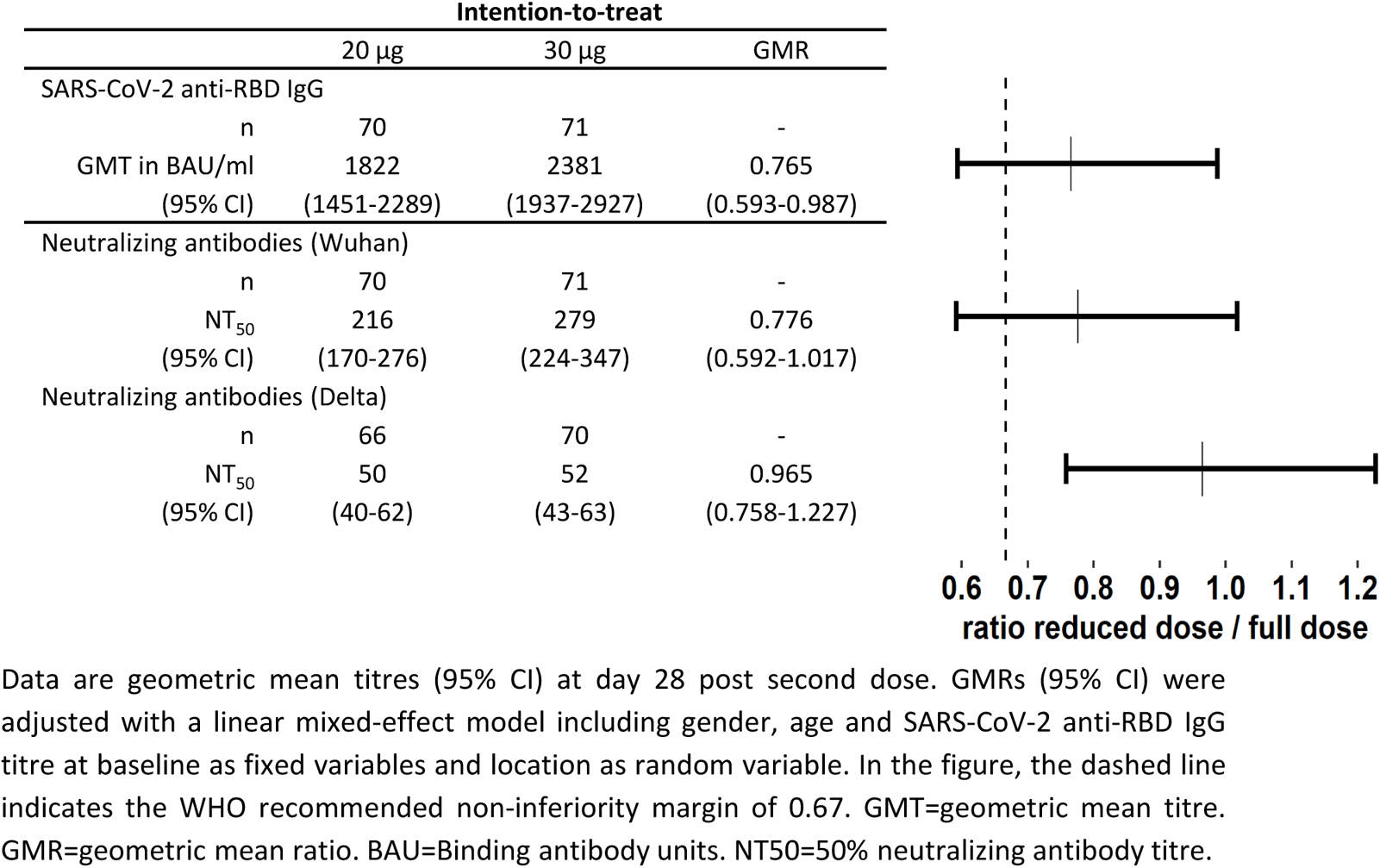
Immune responses by study arm at 28 days post second vaccine dose (Day 49) and non-inferiority analysis in the intention-to-treat cohort.

**S2 Table.**
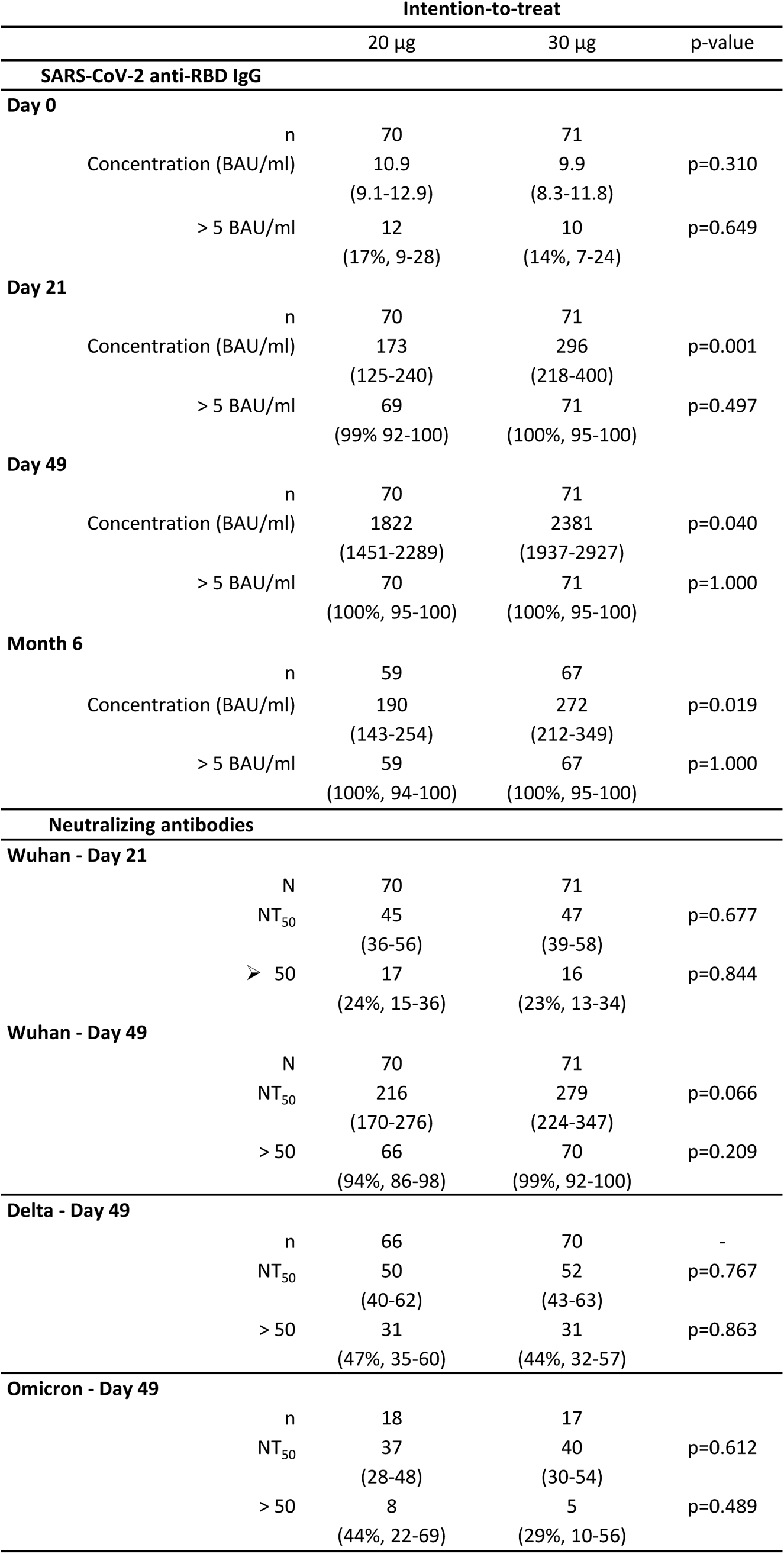
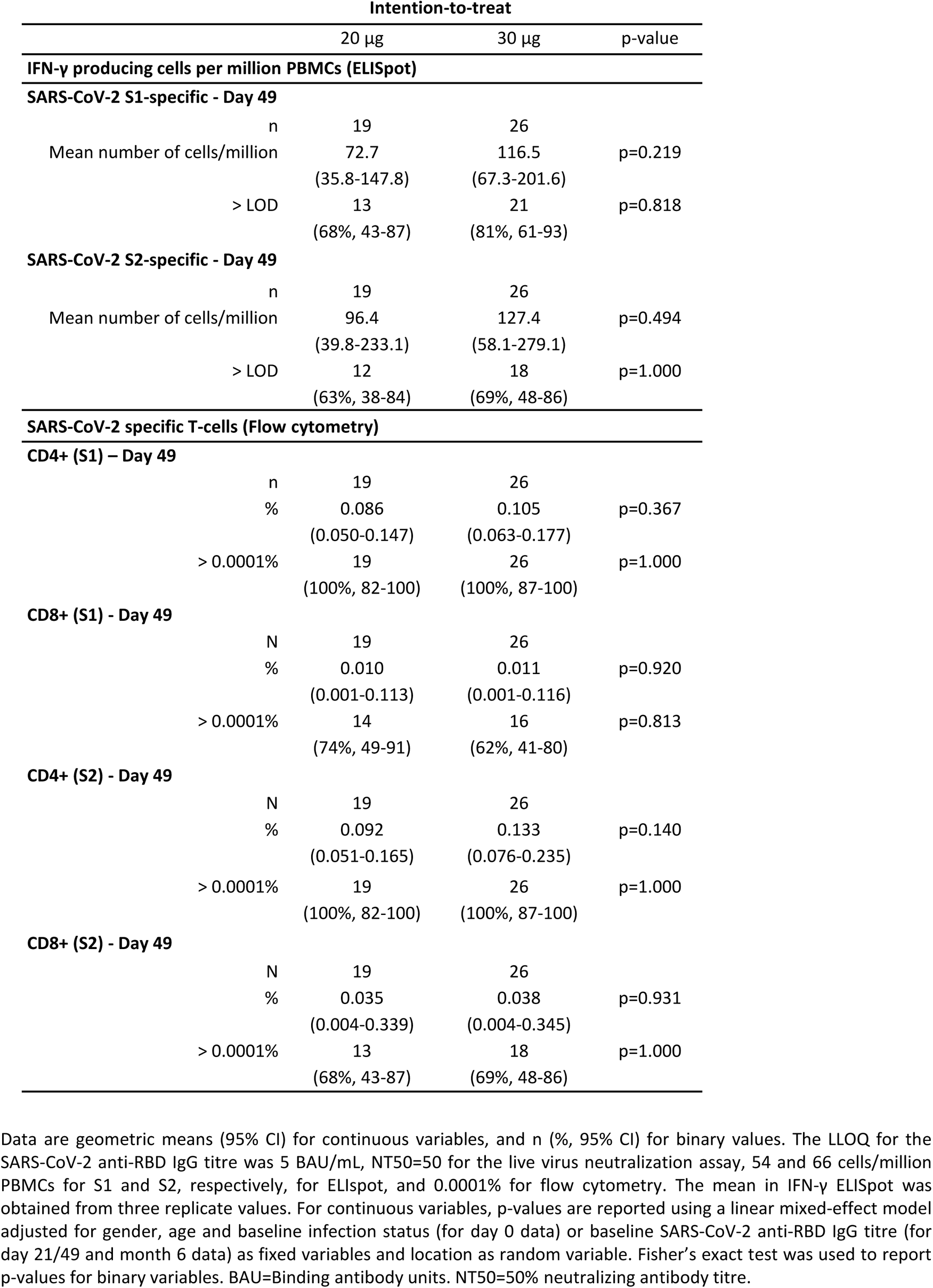
Humoral and cellular responses by study arm in the intention-to-treat cohort at the different time points.

**S2 Fig.**
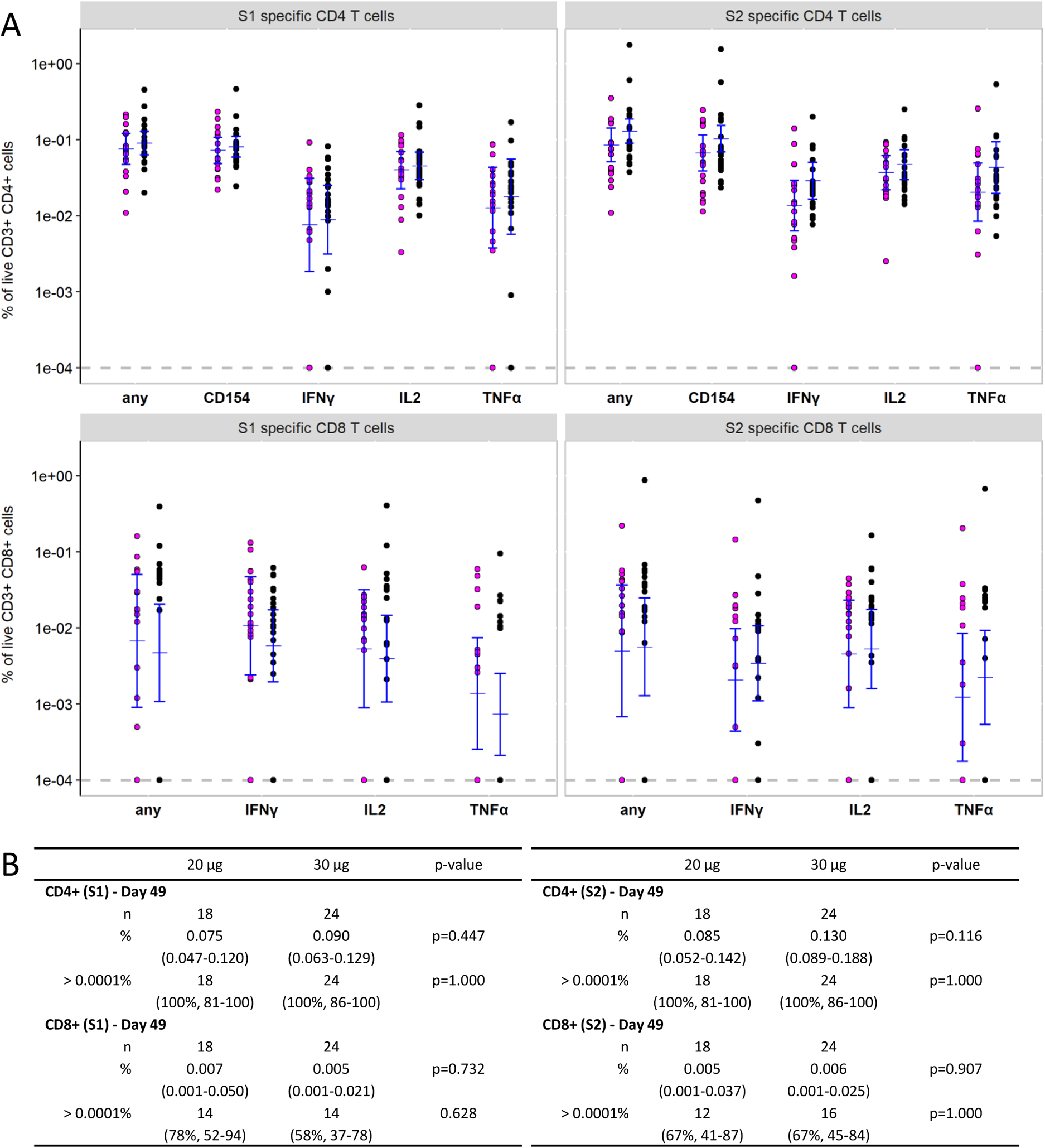
Flow cytometry cellular data in the per-protocol cohort at day 49 (28 days after second dose). SARS-CoV-2 spike protein subunit 1 and 2 (S1 and S2) specific T cell frequencies were measured in 42 infection naïve participants. A. Percentage of CD4+ and CD8+ T cells expressing CD154 (only in CD4), IFN-γ, IL-2, and TNF-α are depicted. Any: percentage of cells positive for at least on the activation markers. A circle represents one test subject; the GMTs (95% CI) are shown in blue. LLOQ was fixed to 0.0001 and represented by a grey dashed line. B. Percentages are given for CD4+ and CD8+ T cells stimulated with S1 Wuhan or S2 Wuhan expressing at least one of the activation markers (corresponding to “any”). For continuous variables, p-values are reported using a linear mixed-effect model adjusted for gender, age and baseline SARS-CoV-2 anti-RBD IgG titre as fixed variables and location as random variable. Fisher’s exact test was used to report p-values for binary variables.

**S3 Table.**
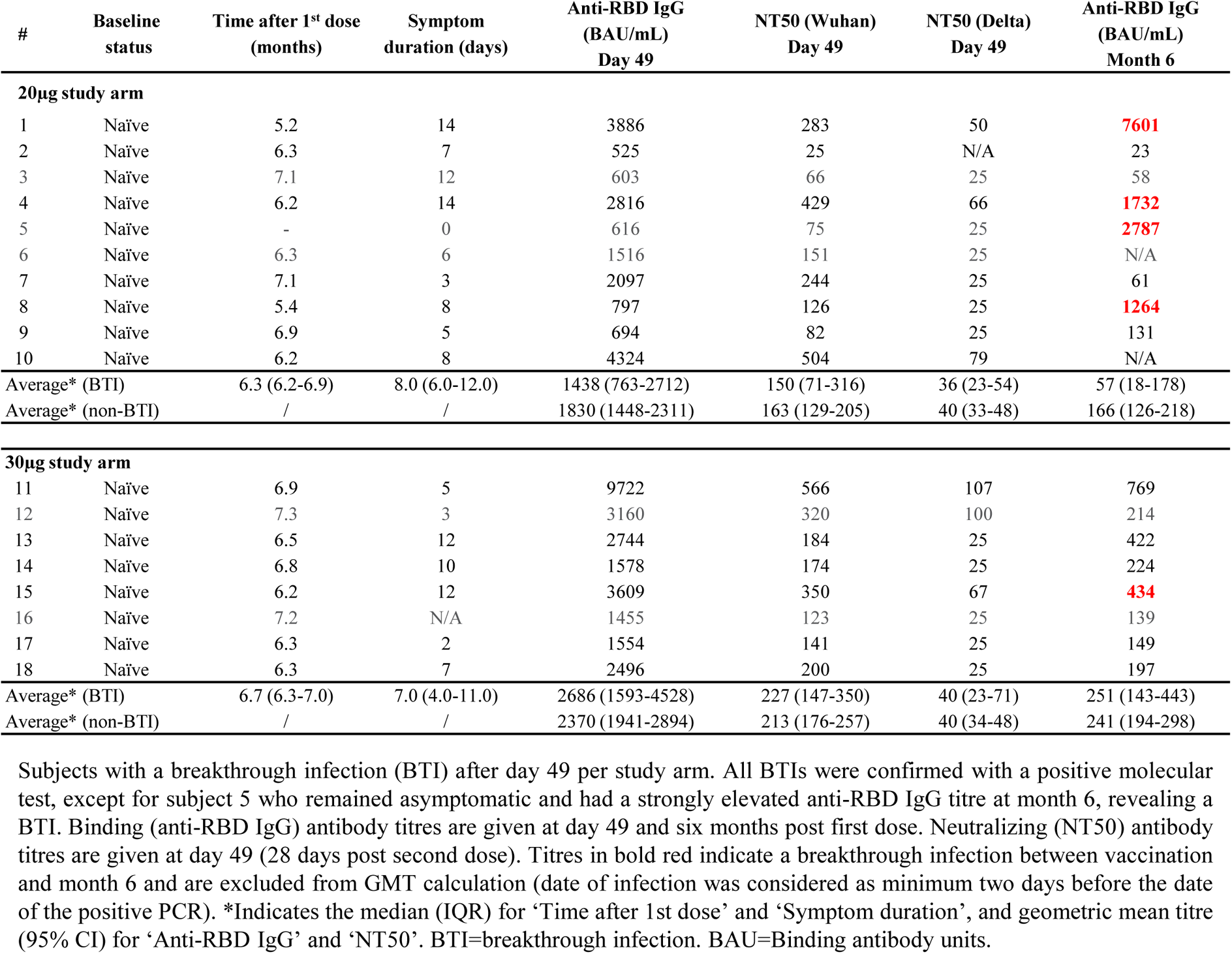
Breakthrough infections

**S4 Table.**
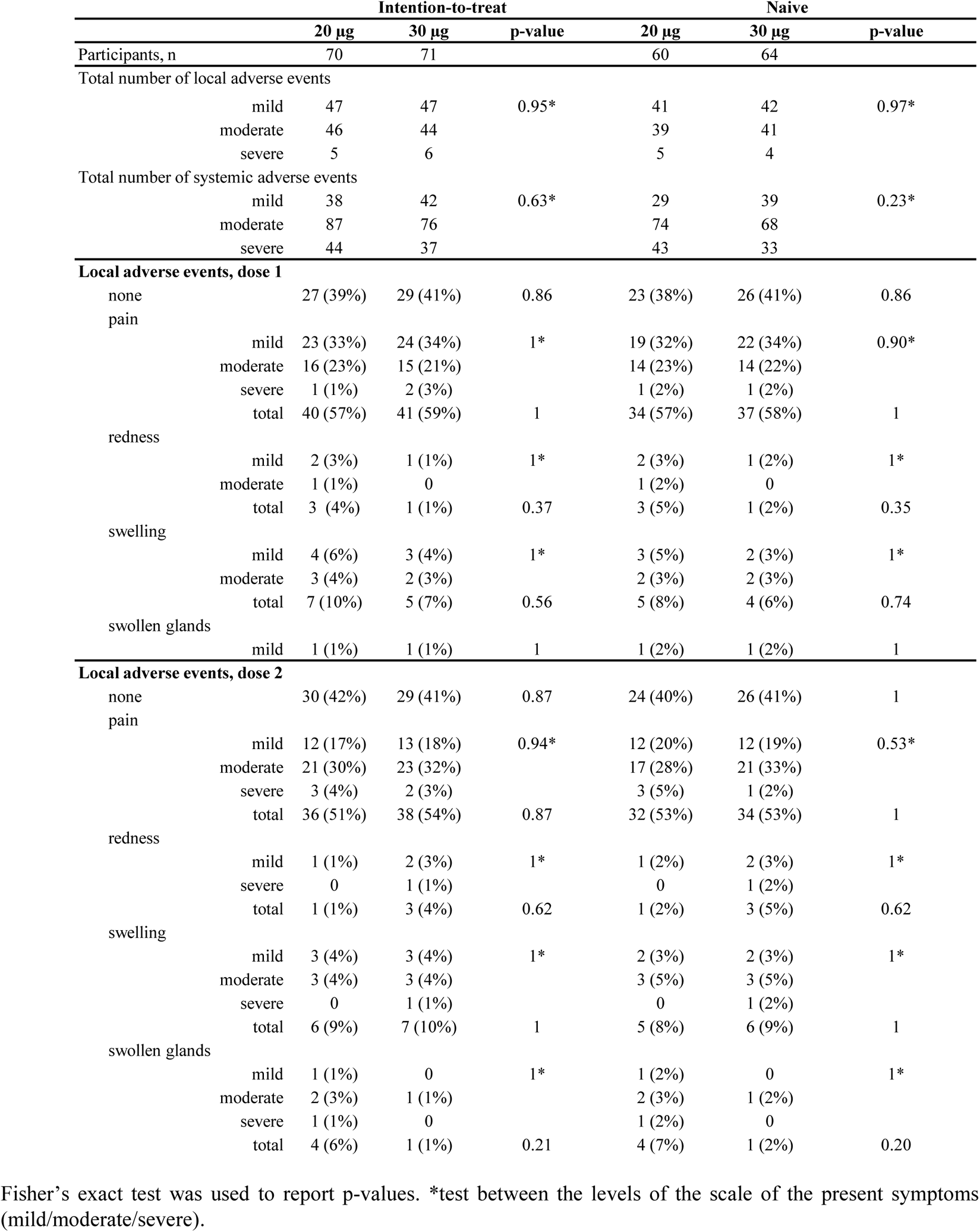
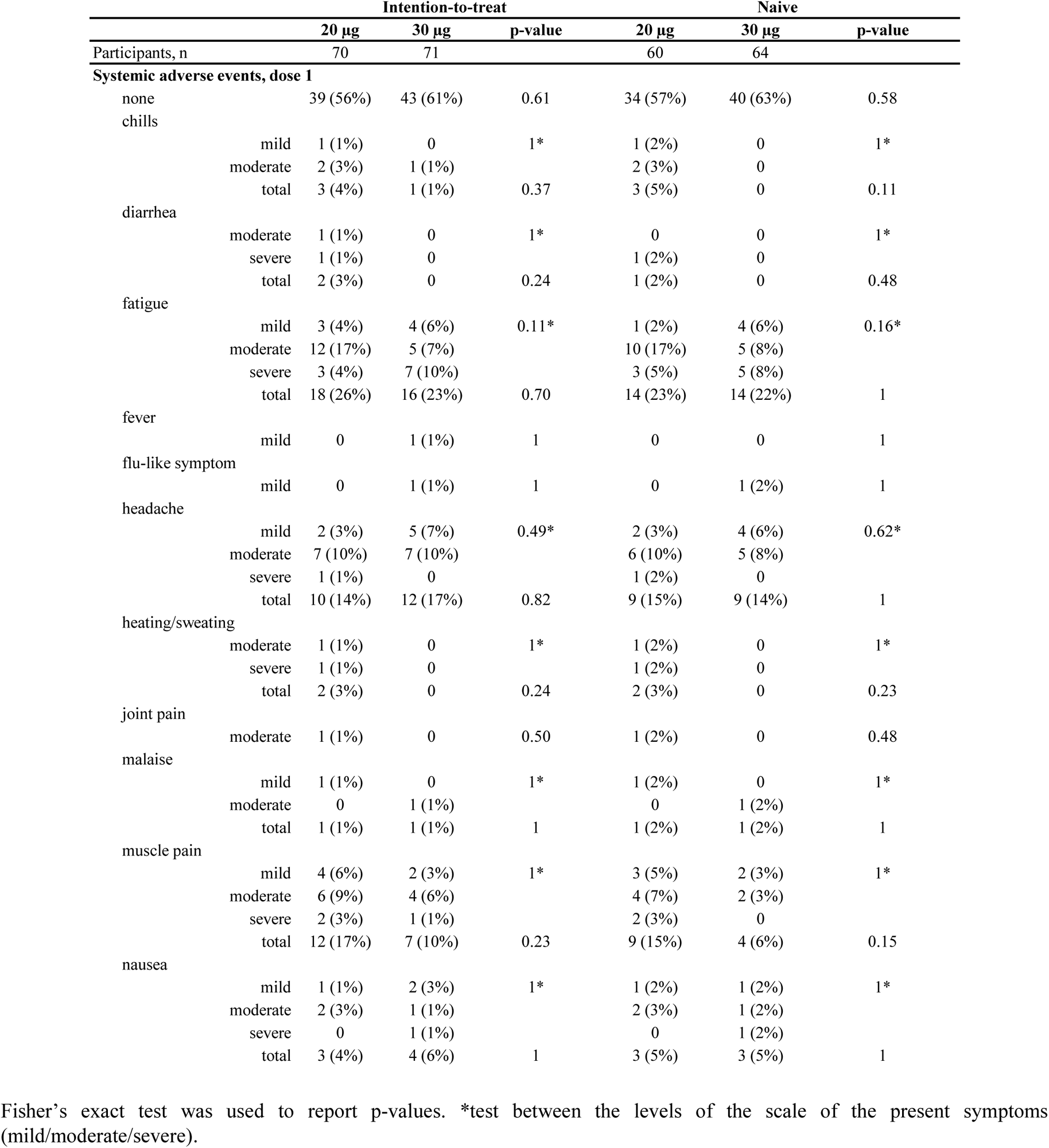
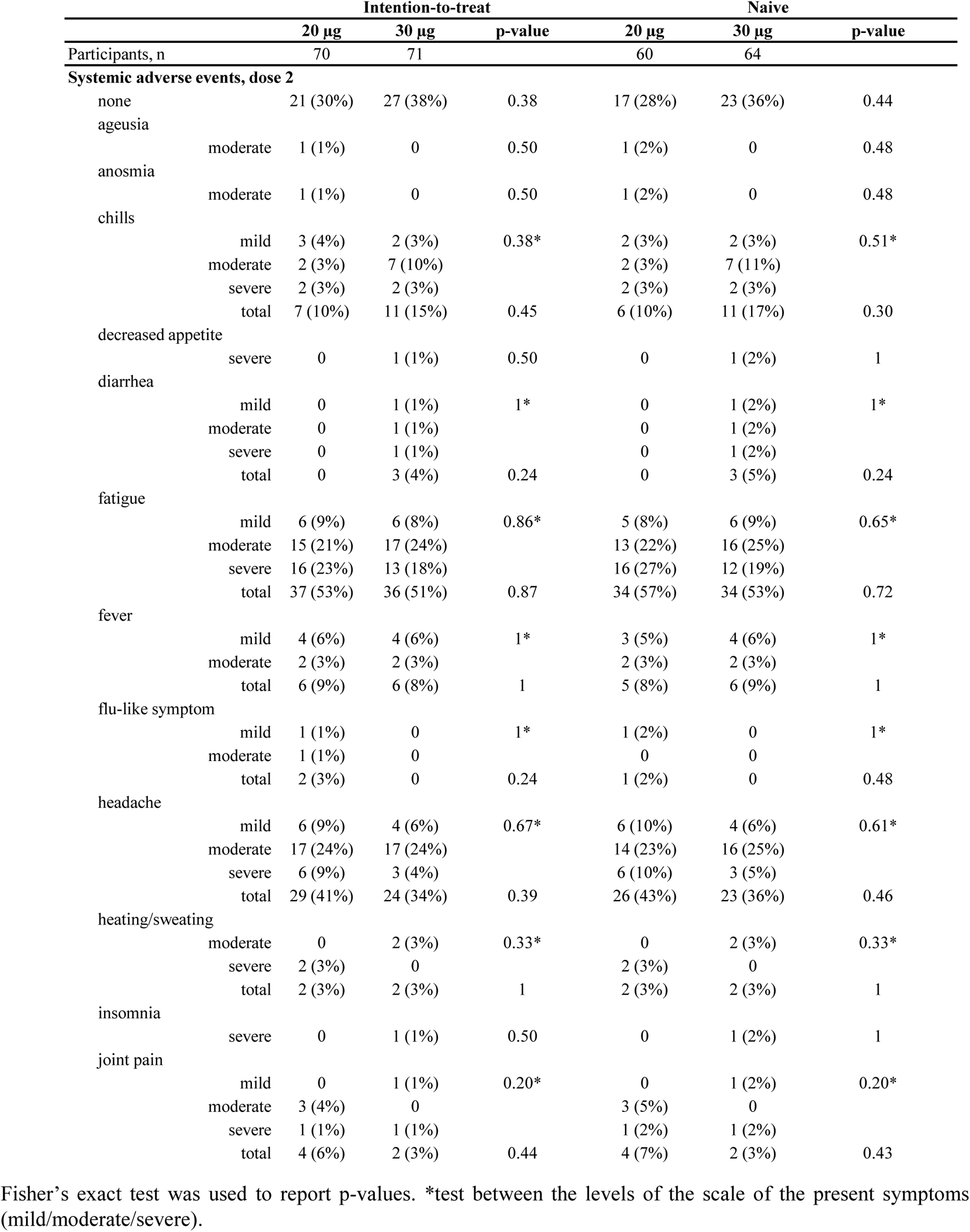
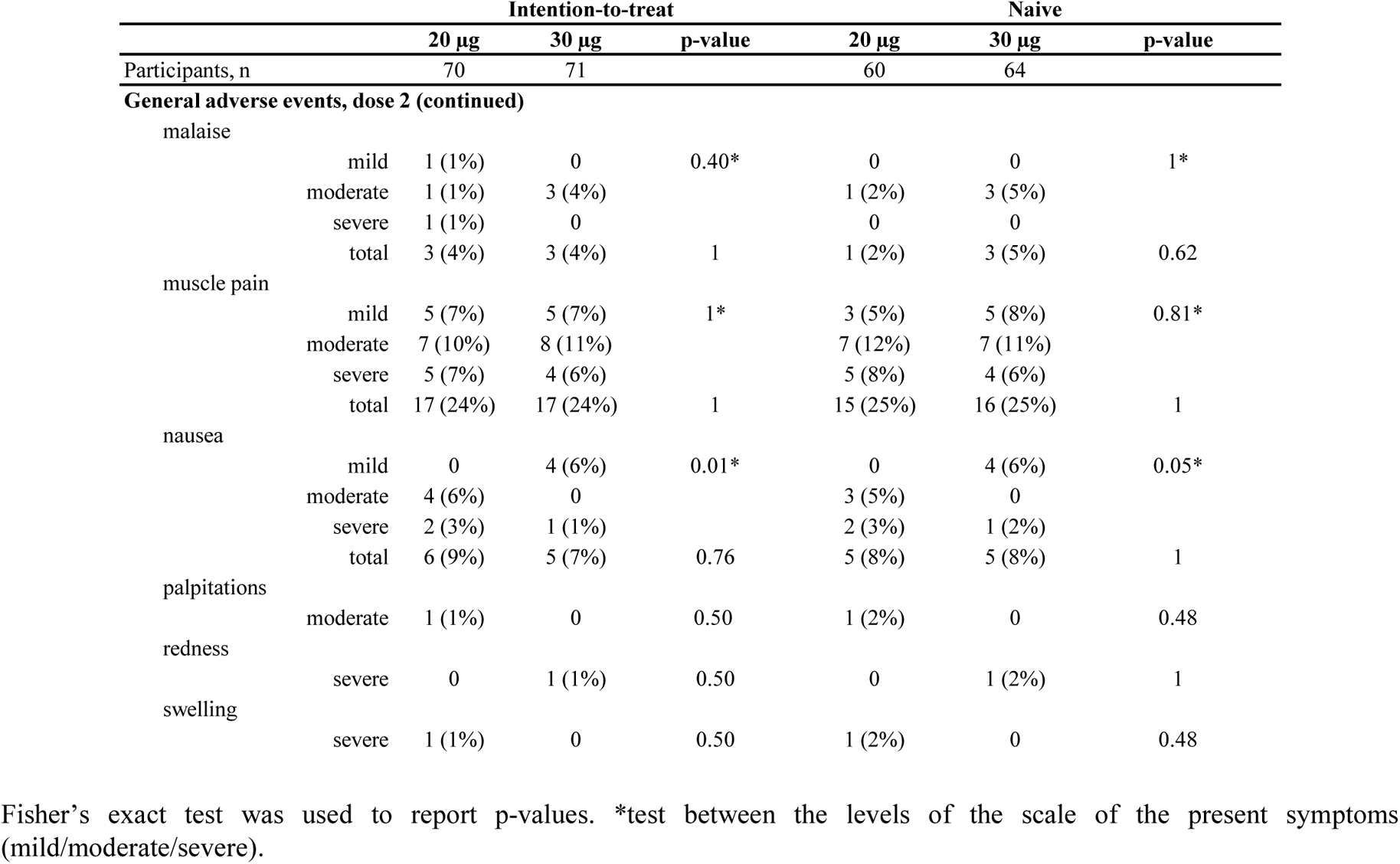
Local and systemic adverse events in intention-to-treat and naïve only cohorts.

**S3 Fig.**
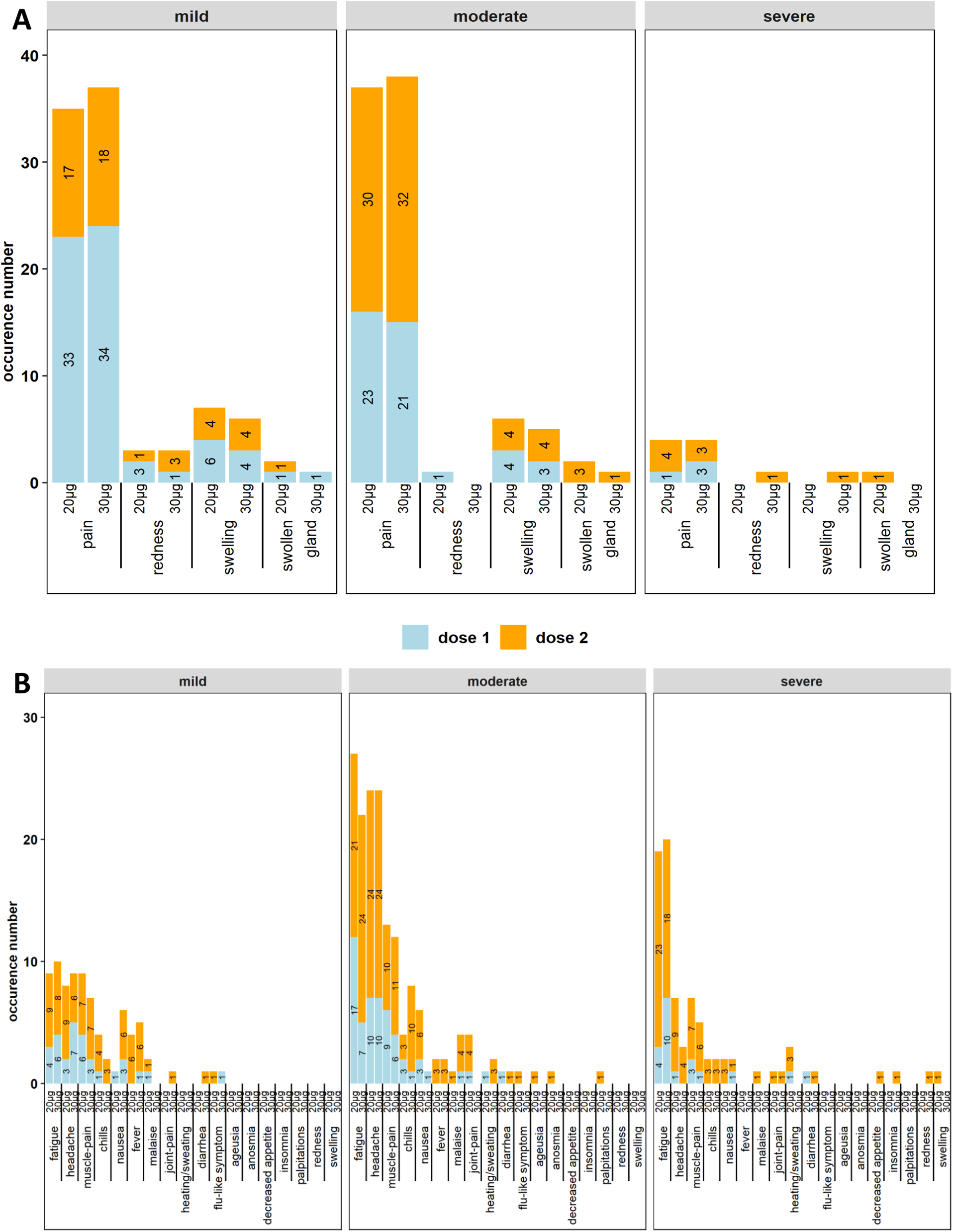
Adverse events. Reported local (A) and systemic (B) adverse events after the first (in blue) and second (in orange) vaccine dose, according to severity (mild/moderate/severe) and by study arm (20µg and 30µg) in the intention-to-treat cohort. Occurrence number (x-axis) and percentage (numbers inside bars) per AE calculated on the cohort are given.

### S1 Appendix: Study protocol

#### 1. General information

This phase IV dose-optimization study will be coordinated by the service Epidemiology of Infectious Diseases and Cancer center (contact: Mieke Goossens, Pieter Pannus) of the scientific directorate of Epidemiology and public health, Sciensano. Laboratory analyses will be performed by the service Immune response (contact: Isabelle Desombere) and Viral diseases (contact: Cyril Barbezange, Isabelle Thomas) of the scientific directorate of Infectious diseases in humans of Sciensano. The study will be executed in collaboration with Mensura EDPB (contact: Marie-Noëlle Schmickler and Mathieu Verbrugghe), the Virology Unit of the Institute of Tropical Medicine Antwerp (contact: Kevin Ariën), the Institute for Medical Immunology of ULB, Campus Erasme, (contact: Arnaud Marchant) and Campus Gosselies (contact: Stanislas Goriely).

#### 2. Background

COVID-19 vaccines are being rolled out in many countries all over the world. Promising efficacy data from phase three vaccination trials are being confirmed with real-world data from countries like Israel and the United Kingdom where large proportions of the population have already been vaccinated (1–3).

Vaccine supply has proven an important limiting factor for the speed of vaccination campaigns. Indeed, vaccine demand largely surpasses production capacity of the different manufacturers. In order to cope with this scarcity, different strategies have been proposed and implemented, including a delayed second dose and a single instead of dual dose for previously infected people. Another possible strategy is a reduction of the vaccine dose for those population groups who generally have better immunologic vaccine responses, which is the subject of this clinical trial (4). Indeed, in this study we will investigate immune responses to a reduced dose of the BNT162b2 mRNA vaccine of BioNTech/Pfizer.

Data from a dose-escalating phase 1 trial in healthy adults 18 to 85 years of age comparing two doses of 10µg, 20µg, 30µg and 100µg indicated that a dose of 30µg of BNT162b2 achieved the best immune response in participants of all ages (5). As a result, a phase2/3 trial was conducted and the vaccine was finally marketed at a dosage of 30µg (6). These immune responses were age dependent, however. While people aged 65-85 years (N=24) had markedly better responses with 30µg as compared to 20µg, this was not the case for people aged 18-55 years (N=24). Indeed, SARS-CoV-2 specific binding and neutralizing antibody titers were even slightly higher in the 20µg group as compared to the 30µg group. We therefore propose to conduct a clinical trial investigating immune responses comparing a 20µg versus 30µg dose of BNT162b2 in a larger cohort of 150 subjects..

#### 3. Objectives and outcomes

We aim to include 150 adults aged 18-55 years from five Mensura EDPB sites. These will be equally randomized in two study arms receiving two doses of:

- Arm 1: 20µg BNT162b2
- Arm 2: 30µg BNT162b2

We aim to include up to 25 previously SARS-CoV-2 infected subjects per arm. The objectives of this study include assessing the immunogenicity (both humoral and cellular), safety and reactogenicity of a reduced vaccine dose.

##### 3.1. PRIMARY OBJECTIVE AND OUTCOME

- The primary objective is to prove non-inferiority of immunogenicity of a reduced BNT162b2 vaccine dose (20µg) versus the reference vaccine dose (30µg).
- The primary outcome is the geometric mean titer (GMT) of binding (IgG) antibodies specific to the receptor binding domain (RBD) of SARS-CoV-2 at four weeks after the second dose.

Data from the literature and from our own PICOV-VAC data indicate strong correlation between titers of RBD binding antibodies and neutralizing antibodies. As methods to measure RBD binding antibodies can be implemented more rapidly than neutralizing antibody assays, we consider that RBD binding antibodies are a good surrogate for neutralizing antibodies and can therefore be used as a primary endpoint for this study.

##### 3.2. SECONDARY OBJECTIVES AND OUTCOMES

- GMT of RBD-specific binding antibodies at the time of the second dose, six months and one year after the first dose.
- GMT of neutralizing antibody titers against wild type and variant SARS-CoV-2 viruses at four weeks after second dose administration.
- Cellular immunity parameters (i.e. Memory B-cell responses, T-cell responses, etc) at all clinically and biologically relevant time points.
- Safety and reactogenicity of the different vaccine regimens as defined by the severity, duration and amount of adverse events experienced after each vaccine dose.

#### 4. Methods

This is a randomized interventional clinical trial in healthy subjects organized at five sites of Mensura EDPB.

##### Inclusion criteria

- Employed by Mensura EDBP (should be an employee at least until the end of the study)
- Aged 18-55 years

##### Exclusion criteria

- Previously vaccinated against COVID-19
- Pregnant/breastfeeding women

##### Discontinuation criteria

All participants retain the right to end their participation in the study at any point in time. Participants who fail to provide the necessary information through the questionnaires, will be discontinued from the study, since any correlation analysis with biological measurements becomes impossible.

The primary objective of the study is to prove non-inferiority of anti-RBD binding antibody titers four weeks after the second dose of the 20µg versus 30µg study arm. If the primary outcome of the study reveals an inferiority of the 20µg dose, participants in the 20µg dose arm will be offered a third, 30µg dose of the study vaccine.

Although no correlate of protective immunity has been validated yet, a relationship between vaccine-induced SARS-CoV-2 antibodies and protection has been proposed (7).

##### Sample size

The primary analysis is a non-inferiority comparison at 28 days after the second dose, for the GMT of antibodies binding to the RBD of SARS-COV-2, comparing the reference dose (30µg BNT162b2) with the reduced dose (20µg BNT162b2).

Currently available data from published and ongoing COVID-19 vaccination trials indicate that:

- RBD-specific antibody titers (GMT, arbitrary units) 28 days after the second dose of BNT162b2 equals 2125. The standard deviation on the log scale (base 10) is 0.27 (PICOV-VAC trial).

The following assumptions are made in the sample size calculations:

- The non-inferiority margin is −0.15 absolute difference of GMT on log scale (base 10), between a reduced dose and regular dose vaccine schedule.
- The standard deviation of GMT on the log10 scale is 0.27 for BNT162b2, based on the currently available data.
- The true difference of GMT on log10 scale is 0.
- A two-sided 2.5% family-wise error rate (at cohort level).

Based on these assumptions, the comparison between both doses will need a minimum of 50 participants (infection naïve) per group to achieve 90% power. In order to be able to better evaluate reactogenicity however, we aim for a larger sample size.

##### Trial randomization

Subject randomization will be done using statistical software (RStudio). The system will randomly allocate the 150 participants across both study arms (20µg and 30µg) irrespective of their previous SARS-CoV-2 infection status. The recruitment and randomization process will aim to have a balanced representation of age and gender across the study arms. Therefore stratified randomization will be used based on age and gender.

In addition, 20 participants will be randomly selected per study arm (40 in total) from which extra (heparinized) blood will be collected for in-depth cellular immunogenicity analyses (see table 1). This random selection will be done using the same statistical software (RStudio).

##### Blinding/unblinding

Everyone involved in the study will be blinded to the study arm allocation of each participant, except for the study nurses administering the vaccine.

When the primary outcome of the study has been reached (28 days after the second dose), an interim analysis will be conducted to determine whether the reduced vaccine dose induces an inferior immune response as compared to the regular dose or not. In case of non-inferiority, there will be no need for unblinding before the end of the trial. In case of inferiority, study participants will be unblinded, informed about which study arm they were allocated to and offered a third dose (of 30µg) of BNT162b2.

##### Study flow

###### Participant information and consent procedure

A research nurse checks the study eligibility criteria and informs the candidates on the possibility to participate in the study. Participant information and consent includes explaining that the national number will be recorded on site by the investigator for possible later data linkage. The participant information and consent will also include that a trusted third party (TTP) will receive and use the national number to link with administrative data. This data linkage is planned to obtain a more complete data set that will be used for the analysis of possible hospitalization and other medical activities billed to the health insurance.

Participant information and consent includes explaining that the email and mobile number from study participants will be recorded in a separate and protected form in the database hosted by the occupational physician. These data are only used to send emails and text messages to the participant for this study.

After having had the possibility to think and discuss about their participation, the participant gives written informed consent and provides the research nurse with their email address and telephone number that they intend to use the next 12 months.

###### Baseline activities

After the participant has given written informed consent, the research nurse completes the Subject ID log with participant’s name, national number and study number. The participant national number is collected and kept on site by the research nurse in the Subject ID log. Each of the participating sites will transfer this local Subject ID log at study closure to the TTP for data linkage, following the procedure detailed in the authorization of the Information Security Committee.

The research nurse creates a new participant in the study eCRF (hosted on LimeSurvey) and enters the date of consent, gender, year of birth, height, weight. They will also register whether the participant has had a past SARS-CoV-2 infection (i.e. positive nasopharyngeal swab, positive serology test).

Once the information is entered by the research nurse in the eCRF, the participant automatically receives an email (subject “Clinical Trial Message” and starting with “Dear Madam, Dear Sir”) with a link to the data entry screens for the patient-reported-outcomes (PROMs). In order to have a second identity check, the participant first enters year of birth and their study number.

The research nurse can view the PROMs of his/her subjects and only exceptionally make corrections upon explicit request by the participant. Each such change will be logged and needs a justification. At all times the research nurse can indicate in the system the early end of study (and stop sending emails to the participant to complete the PROMs), e.g. in case of withdrawal of consent, if known to the research nurse. In case of withdrawal of consent, data already collected will be kept in the study data.

###### Vaccination

Depending on the randomization, participants will either receive two doses of 20µg or two doses of 30µg of the BioNTech/Pfizer SARS-CoV-2 mRNA vaccine, with a three week interval between both doses.

###### Sampling visits

An overview of the different sample types and volumes at each study visit is summarized in table 1. Venous blood will be collected on the day of the first and second dose as well as four weeks after the second dose, and six months and one year after the first dose. Five mL of blood will be collected from all study participants at all study visits. From a random selection of 20 participants per study arm (40 in total), 36mL of heparinized blood will be collected on the day of the first dose as well as four weeks after the second dose and six months after the first dose.

The sampling activity (date, sample types collected) is documented in the eCRF. Samples are handled as detailed in the sampling manual.

One week after each vaccination dose, participants will be invited by email and text message by the research nurse to fill in a questionnaire collecting information on the reactogenicity of the vaccine.

###### Monitoring of adverse reactions

Suspected unexpected serious adverse reactions, serious adverse reactions, and adverse reactions with grade equal or more than 3 will be monitored for the duration of the study period. The study nurse will complete the reporting form (annex 4) and forward this to the study coordinator, who in turn will report this immediately to the Belgian Federal Agency for Medicines and Health Products.

###### End of the trial

The trial will end when the last study participant has been sampled one year after first dose administration.

##### 4.1. DATA MANAGMENT

###### Safety

Based on previously published phase 2 and 3 data of both vaccines that will be used in this study, very few to no serious adverse events are expected (6, 8). Besides vaccination, no safety issues linked with the other study procedures are expected either. As a consequence this is considered a low risk study.

###### Data collection

Laboratory data (samples) and epidemiological data (questionnaire) will be collected as described above. Each participant will receive a unique identifier code at the start of the study that will be used both on the samples and the questionnaires through the whole study period.

###### Laboratory data

Blood samples will be collected as indicated in table 1. Serum will be isolated from serum gel blood collection tubes and will be used for:

- SARS-CoV-2 specific binding antibody quantification: using an *in-house* anti-RBD IgG ELISA.
- Neutralizing antibody capacity measurement against the wild type Wuhan strain and at least one variant of concern: using a virus neutralization assay.

Peripheral Blood Mononuclear Cells (PBMC) will be isolated from heparinized blood on the day of blood draw and will be stored in liquid nitrogen for further analyses. These include:

- Memory B-cell responses: using B-cell ELISpot of flow cytometry
- T-cell responses: using intra-cellular cytokine staining

Samples will be collected at the research sites by a research nurse of Mensura EDPB.

###### Epidemiological data

Socio-demographic characteristics and SARS-CoV-2 initial status (date of last PCR test) will be collected by the research nurse at the time of inclusion and will be pseudonymized (without the identity of the participant but with a unique participant code, see further) transferred to Sciensano after informed consent of the participant was obtained. Additional questionnaires will be filled in by the participants given time points (see Table 1), and will provide information on reactogenicity and severity of the adverse events. All questionnaires will be completed through a secured online application (see further).

###### Data flow and management

Blood collection tubes as well as labelling stickers will be provided by Sciensano. For the entirety of the study, material will be provided in batch and a transport will be organized between Sciensano and the research sites. After collection, samples will be handled following the appropriate SOP.

Epidemiological and laboratory data will be linked via a unique code assigned to each participant. This code will start with the first letters of the research site location followed by a three digit number (e.g. “BRU_001” for the first participant, “BRU_002” for the second one, and so forth).

A sticker labelled with this code will be affixed by the research nurse on each tube at the time of sampling, and the same corresponding code will be entered in each questionnaire, enabling the link for data analysis. For the planned days, the labels will also include an identifier of this point (e.g. “D00”, “D28” etc). This unique code must stay the same during all the duration of follow-up and a list of all participants and their assigned codes will be kept in a secure and protected way by the occupational physician.

Questionnaires are filled in online through LimeSurvey. Before sampling takes place, an email will be sent to the participant containing a questionnaire link and the unique participant code. It will take around five minutes for the participant to fill in the questionnaire. If the questionnaire is not filled in yet at the moment of sampling, it will be filled in together with the research nurse. A confirmation email will be sent to the research nurse when a questionnaire is correctly received. LimeSurvey is a web application running on a server located within Sciensano’s datacenter. The application and its associated database are located on the same server. It means there are no data located outside Sciensano. Access to the administration of the LimeSurvey application is restricted to a limited number of people involved in the administrative management of the survey, who are authenticated with a username and password. All files will be kept on the Sciensano SQL server with restricted access.

Login to the shared server is password controlled. Each user/investigator or research nurse will receive a personal login name and password and will have a specific role which has predefined restrictions on what is allowed on the server. Furthermore, users will only be able to see data of subjects of their own site. Any activity in the software is traced and transparent via log files.

Direct access to all study records, including source documents and the eCRF will be granted to authorized representatives from the Sponsor, host institution and the regulatory authorities to permit study-related monitoring, audits and inspections. The Data Manager will review the eCRFs to make certain that all items have been completed. Incorrect or inappropriate entries in the eCRFs will be returned to the research nurse for correction (data queries), if not already captured using automated data entry checks. Subject privacy must be respected at all times, in accordance to GDPR, GCP and all other applicable local regulations. The investigator/study team should immediately notify the sponsor if he or she has been contacted by a regulatory agency concerning an upcoming inspection.

The data capture software analysis, development and testing will be performed according to the procedures of the Sponsor. Quality check of data management and data entry will be performed in accordance with the standard operating procedures of the Sponsor.

###### Data analysis

Analysis of all data collected within this study will be performed by Sciensano. Questionnaire responses will always be coded. All analyses will be performed in R (R Core Team (2018), available from https://www.R-project.org/), STATA or SAS.

Meaningful laboratory results of each participant will be communicated to the research nurse who will be able to communicate them to the participants. This includes the anti-SARS-CoV-2 antibody status at the primary endpoint and at the end of the study.

As already mentioned, these tests are for research and not diagnostic purposes, and depending on lab capacity, there will be a delay in the communication of the results. The global results of the study will be communicated to each participating center at the end of the study in the form of a report and/or presentations.

###### Statistical analyses

The analysis described here will be coordinated by the Sponsor. Any later analyses on the linked data will also be conducted by the Sponsor.

All of the participants who meet the eligibility criteria will be included in the main analysis. Descriptive analyses will be used to report participant characteristics. The level of antibody titers in each of the study arms will be analyzed with the Kruskal-Wallis test, while participant demographics (e.g. age, gender) will be taken into account (confounders). Furthermore, risk factors and correlations with antibody detectability will be evaluated. P-values below 0.05 will be considered statistically significant.

#### 5. Ethics and Privacy Protection

##### 5.1. INDEPENDENT ETHICS COMMITTEE AND INFORMED CONSENT

Prior to study start, this protocol will be reviewed and approved by an Independent Ethics Committee (IEC). Written, dated and signed informed consent has to be obtained from each eligible subject prior to inclusion in the study. A sample Participant Information/Informed Consent Form has been prepared (see appendix).

##### 5.2. DATA LINKAGE

An electronic case report form (eCRF) will be used for data collection. Subject confidentiality will be maintained at all times, within the legal constraints. The exported participant reported outcome measures (PROMs) dataset will contain pseudonymized participant data. A separate instrument (“form”) with separate restricted access will contain direct identifiable subject data (subject’s email address and mobile phone number of the participant) such that emails and additional reminders by text message can be sent to the subject. Separately from the study database, the local investigator will store on their Mensura EDPB facility a Subject ID log (e.g. in a spreadsheet with restricted access, GDPR compliant) containing the national number of the participant and the unique participant number for a possible later data linkage by a trusted third party (TTP), f.e. eHealth. The linkage will follow a procedure approved by the competent chamber of the Information Security Committee.

After the Ethics Committee has approved this protocol, the Sponsor will introduce a request to the competent chamber of the Information Security Committee for the linkage of the identifiable study data including participant reported outcome measures (PROMs), sickness fund (IMA/AIM) financial data (RIZIV/INAMI expenses). Only after the approval of this request, the parties will start data linkage. The principle investigator will guarantee the data protection.

#### 6. Organization of the research project

##### 6.1. TIMELINE

The anticipated start date of the study is May 3^rd^ 2021. The end of the trial is anticipated to be the beginning of June 2022.

##### 6.2. STUDY PARTIES AND RESPONSIBILITIES

###### Study sponsor: Sciensano

Study contact and principle investigator is Dr. Maria Goossens, Sciensano. The sponsor has in-house expertise in the lab tests.

The sponsor has in-house expertise with LimeSurvey for both the eCRFs and the collection of PROMs. Data will be captured on a secured server.

The biostatistician of the Sponsor will perform the study data analysis.

###### The independent Ethic committee: ULB Erasme

The current epidemic context justifies the rapid implementation of this study, with the aim of improving the care of the population and contributing to a better control of the pandemic.

###### Trusted Third Party (TTP): e-Health platform

The national number of participants is stored on site in the subject ID log.

The PROMs will be linked by the TTP using the national number with the invoices of medical acts in the IMA database, after approval by the competent chamber of the Information Security Committee.

###### Participating sites and local investigators

The local investigator, the occupational physician, will collect written informed consent from all participants. In order to facilitate the logistics of participant inclusion, a form to detail participants included will be provided to the investigators. This Subject ID log (appendix) is to be kept by the investigator on site and contains the national number. A copy of the study Subject ID log with the national number and study subject ID is to be provided to the TTP for data linkage.

##### 6.3. RESOURCES

- Purchase, preparation and transport of material for the study will be foreseen by Sciensano.
- Mensura EDPB will organize the research sites through the occupational physician.
- Lab analyses will be performed at Sciensano, ULB and ITG.
- PBMC preparations will be done by Sciensano, ULB and ITG.
- Epidemiological and laboratory data will be analyzed by the Sciensano researchers.
- A budget of 203 600 € was estimated and approved.

#### 7. Risk and benefits for participants

##### 7.1. RISKS

The risks for participants are small, they include side effects of the sampling procedure and vaccination.

- A blood draw can sometimes result in a local hematoma, and rarely in vagal discomfort. Epistaxis can occur in people taking anti-coagulants.
- The most common side effects after BNT162b2 vaccination are usually mild or moderate and resolve within a few days. They include pain and swelling at the injection site, tiredness, headache, muscle and joint pain, chills and fever. They affect about 10% of vaccinated subjects (6).
- Allergic reactions in response to the vaccines do occur, which are severe (anaphylaxis) in very few cases. As for all vaccines, the BNT162b2 vaccine will be given under close supervision with appropriate medical treatment available.

Participants will be clearly informed about these risks, that are minimized due to the expertise of the persons in charge of collecting the samples.

##### 7.2. BENEFITS

In general, the study will provide insight in the possibility of reducing the vaccine dosage required to achieve acceptable immune responses. This might help alleviate the current vaccine scarcity and speed up vaccination campaigns.

In particular, study participants will have the possibility to be vaccinated significantly earlier than foreseen by the Belgian national vaccination campaign, since they all belong to low priority groups. Secondly, those participants receiving a lower dose of the vaccine are expected to experience fewer and less severe side effects than they would with a full dose vaccination (5, 9). Finally, if immune responses in the lower dose arm turn out to be inferior to the regular dose arm, these participants will be offered a third, regular dose to boost their immunity.

##### 7.3. CONFIDENTIALITY

At the researcher level, sample results and questionnaires will be pseudonymized via an individual code attributed to each participant. None of the researchers who analyze the data will be involved in participant data collection, nor in the care of COVID-19 participants. Test results will be communicated by the laboratory to the occupational physician of Mensura EDPB or the research nurse by the individual code attributed to each participant. At all times, the occupational physician and the research nurse are the only ones with access to the identity of the participants.

##### 7.4. BIOLOGICAL SPECIMENS

All the samples, except the PBMCs, collected during the study will be stored in the biobank of Sciensano. Residues remaining after the analyses will be kept for a duration of maximum 10 years if the participant has given consent that the samples can be kept for further research. After 10 years, they will be destroyed. Specimens will be stored in the biobank “Biobank - Sciensano WD12 infectieziekten mens COVID-19” approved by the ‘Commissie voor Medische Ethiek UZ Gent’ on 17 April 2020 (internal reference: EC 037-2020/mf, appendix 3) with registration number assigned by the FAGG/AFMPS: BB200027.

The PBMC samples will be stored in the Biobank from the laboratory where the preparation took place (Sciensano, ITG or ULB):

- ITM Biobank approved by the ‘Commissie voor Medische Ethiek Universitair Ziekenhuis Antwerpen en de Universiteit van Antwerpen” on 8 April 2019 (internal reference: 19/13/168) with registration number assigned by the FAGG/AFMPS: BB190041
- “Biobanque de l’Institut d’Immunologie Médicale (IMI)” approved by the ‘le Comité d’Ethique hospital-facultaire Erasme-ULB’ on 4 May 2020 with registration number assigned by the FAGG/AFMPS: B2020/002
- Biobank - Sciensano WD12 infectieziekten mens COVID-19” approved by the ‘Commissie voor Medische Ethiek UZ Gent’ on 17 April 2020 (internal reference: EC 037-2020/mf, appendix 3) with registration number assigned by the FAGG/AFMPS: BB200027.

##### 7.5. INFORMED CONSENT

Information on the study will be provided by Mensura EDPB and informed consent will be obtained from all participants. The informed consent can be found in appendix 1.

##### 7.6. ETHICS COMMITTEE

The study will be conducted in compliance with the principles of the Declaration of Helsinki (2008) and all of the applicable regulatory requirements.

##### 7.7. PROTOCOL AMENDEMENTS

Any substantial change, clarification, or addition to this protocol requires a written protocol amendment, and this must be approved by the Sponsor before the change or addition can be considered effective. In addition, the IEC should be notified and formal approval by the IEC should be obtained as applicable, before the substantial amendment can be implemented. The substantial amendment will reference the protocol by title and version date and must be signed by the principle investigator prior to initiating the change. Once approved, an amendment becomes an integral part of the protocol.

##### 7.8. INSURANCE

During their participation in the clinical investigation the participants will be insured as defined by legal requirements. An insurance with no fault responsibility has been foreseen by the sponsor in accordance with the Belgian law concerning experiments on humans, 7 May 2004.

#### 8. Appendices

**8.1. APPENDIX 1: INFORMATION BROCHURE AND INFORMED CONSENT**

**8.2. APPENDIX 2: QUESTIONNAIRE**

**8.3. APPENDIX 3: APPROVAL BIOBANK**

**8.4. APPENDIX 4: SUSAR FORM**

### 10. Tables

**Table 1:**
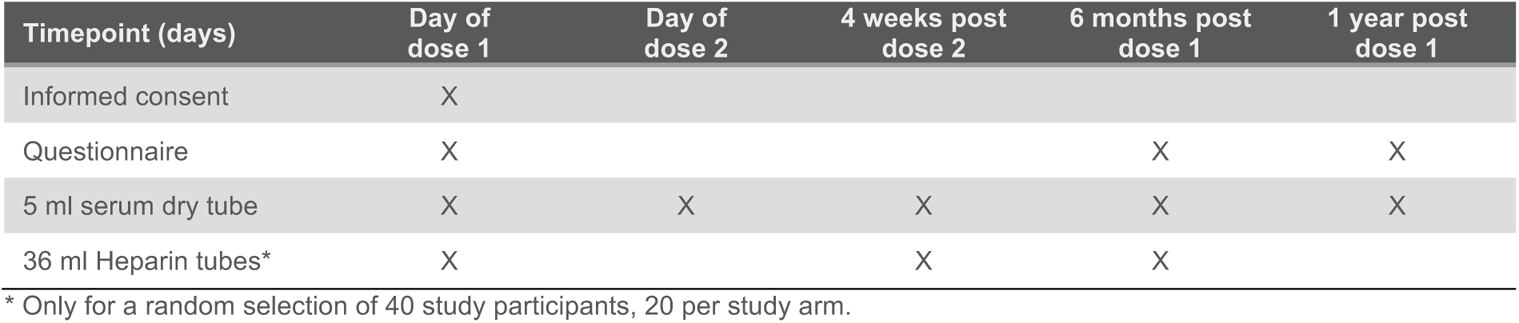
Timetable of data and sample collection.

## S2 Appendix: Supplementary methods

### SARS-CoV-2 Specific Binding Antibodies

#### Enzyme-linked immunosorbent assay

Binding antibodies at baseline and after vaccination were assessed using an enzyme-linked immunosorbent assay (ELISA) for the quantitative detection of IgG-class antibodies to RBD (Receptor Binding Domain, Wuhan strain) (Wantai SARS-CoV-2 IgG ELISA (Quantitative); CE-marked; WS-1396; Beijing Wantai Biological Pharmacy Enterprise Co., Ltd, China). For quantification of antibodies, diluted serum samples (1/10, 1/100, 1/400, 1/1600 and 1/6400) were tested with an internal standard, calibrated against NIBSC 20/136 (First WHO International Standard Anti-SARS-CoV-2 Immunoglobulin), and an external positive control sample included on each plate. Diluted samples were incubated (37°C, 30 min.) with pre-coated micro wells and washed five times. Next, plates were incubated (37°C, 30 min) with horseradish peroxidase (HRP)-conjugated anti-human IgG antibodies and washed five times before adding a TMB and urea peroxide solution for 15 min (37°C, dark). After incubation, a stop solution (0.5 M H2SO4) was added and optical density (OD) was measured at 450 nm using a microplate reader. Net OD values were converted to arbitrary IgG units per ml by interpolation from a point-by-point plot fitted with the standard concentrations and net OD values (correlation coefficient R2≥0.9801), using GraphPad Prism version 9.0.0 for Windows (GraphPad Software, San Diego, California USA) and exported to Microsoft Excel. Antibody measurements were adjusted for any sample dilution, converted to international units per ml (IU/ml) and reported as such. Lower limit of quantification (LLQ) was 5 IU/ml. Clinical performance characteristics of the assay, evaluated in 69 PCR-confirmed COVID-19 patients (comprising mild and severe clinical outcomes, ≥ 15 days post onset of symptoms) and 167 pre-pandemic sera, resulted in a specificity of 100% (95% CI 97,75-100) at a sensitivity of 100% (95% CI 94,73-100) for a cut-off of 6 IU/ml.

#### Multiplex Immunoassay (Luminex)

Antibody responses at baseline were tested with an in house multiplex immunoassay (MIA). In this test, IgG antibodies to SARS-CoV-2 antigens RBD, S1, S2 and N (Wuhan strain) were measured simultaneously in one assay run. In short, purified antigens RBD (cat n° PX-COV-P046, ProteoGenix, Schiltigheim, France), S1 (cat n° PHA002, Sanyou Biopharmaceuticals, China), S2 (cat n° 40590-V08H1, Sino Biological, China) and N (cat n° PNA006, Sanyou Biopharmaceuticals, China) were coupled covalently to distinct color-coded activated carboxylated beads (Luminex, Austin, Texas, USA). Diluted serum samples (1/100, 1/400, 1/1600 and 1/6400, 1/25600) were measured with the international standard (NIBSC 20/136; first WHO International Standard Anti-SARS-CoV-2 Immunoglobulin), control sera and blanks included on each plate and MFI was converted to IU/mL by interpolation from a five-parameter logistic standard curve.

### SARS-CoV-2 Neutralizing Antibodies

Serial dilutions of heat-inactivated serum (1/50-1/25600 in EMEM supplemented with 2mM L-glutamine, 100U/ml - 100μg/ml of Penicillin-Streptomycin and 2% fetal bovine serum) were incubated during 1h (37°C, 7% CO2) with 3xTCID100 of a wild type Wuhan strain (2019-nCoV-Italy-INMI1, reference 008V-03893), the B.1.617.2 Delta variant (83DJ-1) and the BA.1 Omicron variant of SARS-CoV-2, in parallel. Sample-virus mixtures and virus/cell controls were added to Vero cells (18.000 cells/well) in a 96-well plate and incubated for five days (37°C, 7% CO2). The cytopathic effect caused by viral growth was scored microscopically. The Reed-Muench method was used to calculate the neutralizing Ab titer that reduced the number of infected wells by 50% (NT50), which was used as a proxy for the neutralizing Ab concentration in the sample (1–3).

### SARS-CoV-2 specific cellular responses

#### Enzyme-linked immunosorbent spot

Spike-specific cellular responses were tested after vaccination using the Human IFN-γ ELISpot kit (3420-2H) from Mabtech (Stockholm, Sweden) following the recommended protocol. Plates (MAIPSWU10, Mabtech, Stockholm, Sweden) were activated for 15 sec with 50µl of 70% ethanol, and washed with distilled water. Plates were then coated with human IFN-γ antibody (15 µg/ml) overnight at 4°C, washed and blocked with 200µl of Roswell Park Memorial Institute (RPMI) containing 10% fetal bovine serum (FBS) for at least two hours. Next, triplicates of 250 000 PBMC were stimulated in the presence or absence of PepMix SARS-CoV-2 spike glycoprotein peptide pools (SUB1-SUB2, JPT, Berlin, Germany) at 1µg/ml and incubated for 20 hours in a 37°C humidified incubator with 5% CO_2_. After incubation, the plates were washed and incubated with the human biotinylated IFN-γ detection antibody (1µg/ml) for 2 hours, washed and the streptavidin–Horseradish Peroxidase (streptavidin-HRP) diluted at 1/750 in PBS-0,5% FBS was added for one hour. 3,3’,5,5’-Tetramethylbenzidine substrate was added for minimum 10 min at room temperature. Wells were then washed with distilled water and air-dried. Spot were counted with an ELISpot reader (AID Autoimmun Diagnostika GmbH, Straßberg, Germany), mean values of triplicates were considered for S1 and S2 and expressed per million PBMCs after subtracting the mean of the triplicates of the unstimulated condition. The limit of detection is defined as the mean + 2 SD from naïve subjects at baseline (D0), corresponding to 54 and 66 cells per million PBMCs for S1 and S2, respectively. Data points equal to 0 were attributed the value 1 before transformation.

#### Flow cytometry

Cells were stimulated in 96-well round-bottom plates with 1 × 10^6^ PBMCs in RPMI 1640 medium (Lonza, Basel, Switzerland) supplemented with 10% heat-inactivated FBS (Sigma-Aldrich, Kawasaki, Japan), penicillin/streptomycin, amino acids and PepMix SARS-CoV-2 spike glycoprotein peptide pools (SUB1-SUB2, JPT, Berlin, Germany) in the presence of 1µg/mL purified anti-CD28 antibody (clone CD28.2, BD Biosciences, New Jersey, USA). Both peptide pools were used at 1µg/ml per peptide. Incubation was performed at 37°, 5% CO_2_ for 6 hours with 10µg/ml brefeldin A (Sigma-Aldrich, Kawasaki, Japan) added after 90 min. After stimulation, Live/Dead fixable red stain (ThermoFisher, Massachusetts, USA) was used to exclude dead cells and the staining of surface antigens was carried out for 20 min with the following fluorochrome-conjugated antibodies: CD3 BV711 (UCHT-1; BD), anti-CD8 PeCy7 (RPA-T8; BD), CD4 HV450 (RPA-T4; BD). Fixation and permeabilization were performed with Cytofix/Cytoperm (BD) and intracellular staining was carried out for 30 min: IFN-γ FITC (RPA-T8; BD), IL2-APC (MQ1-17H12; BD), TNF-AF700 (Mab11; BD), CD154 APC-Cy7 (TRAP-1; BD). Cells stimulated with 1mg/ml Staphylococcus enterotoxin (SEB; Sigma-Aldrich) served as positive controls and unstimulated cells only contained anti-CD28. Samples were acquired on a BD LSRFortessa flow cytometer and analyzed with FlowJo v9. The proportion of cells producing cytokines was determined by subtracting the expression levels/production levels in the unstimulated wells, from the peptide stimulated wells.

