## Appendix for "Safety and immunogenicity of a reduced dose of the BNT162b2 mRNA COVID-19 vaccine (REDU-VAC): a single blind, randomized, non-inferiority trial"

### SUPPLEMENTARY MATERIAL


| | page |
|---|---|
| **S1 Fig. Decision tree to determine previous SARS-CoV-2 infection status at baseline.** | 2 |
| **S1 Table. Immune responses by study arm at 28 days post second vaccine dose (Day 49) and non-inferiority analysis in the intention-to-treat cohort.** | 3 |
| **S2 Table. Humoral and cellular responses at the different time points by study arm in the intention-to-treat cohort.** | 4-5 |
| **S2 Fig. Flow cytometry cellular data in the per-protocol cohort at day 49 (28 days after second dose).** | 6 |
| **S3 Table. Breakthrough infections.** | 7 |
| **S4 Table. Local and systemic adverse events in intention-to-treat and per-protocol cohorts.** | 8-11 |
| **S3 Fig. Adverse events.** Reported local (A) and systemic (B) adverse events after the first and second vaccine dose, according to severity (mild/moderate/severe) and by study arm (20µg and 30µg) in the intention-to-treat cohort. | 12 |
| **S1 Appendix: Study protocol** | 13-24 |
| **S2 Appendix: Methods** | 25 |

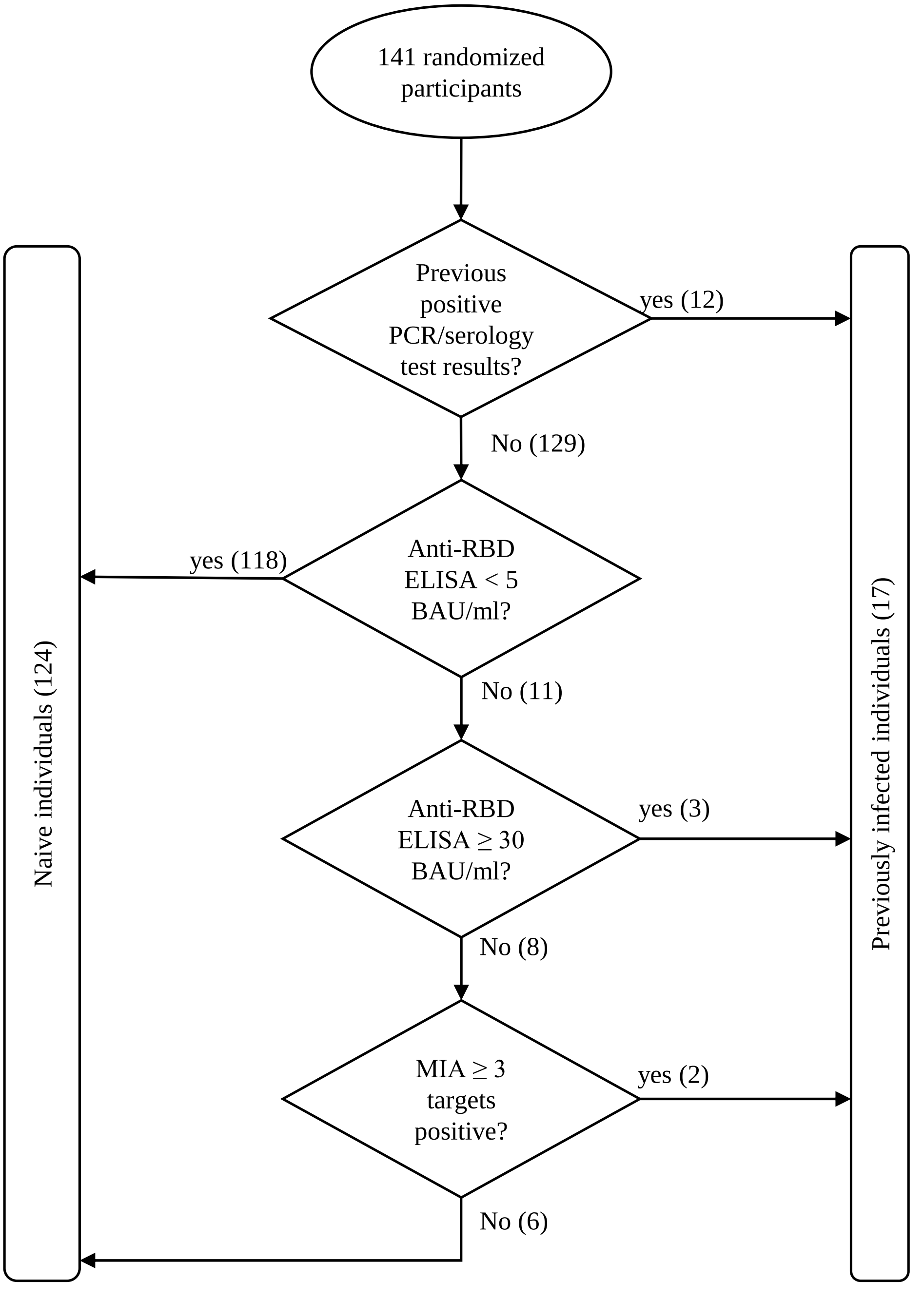


**S1 Fig. Decision tree to determine previous SARS-CoV-2 infection status at baseline**. RBD=SARS-CoV-2 receptor binding domain, ELISA=Enzyme linked immunosorbent assay, BAU=binding antibody units, MIA=multiplex immunoassay.

**S1 Table. Immune responses by study arm at 28 days post second vaccine dose (Day 49) and non-inferiority analysis in the intention-to-treat cohort.**

| | Intention-to-treat | | |
|---|---|---|---|
| | 20 µg | 30 µg | GMR |
| SARS-CoV-2 anti-RBD IgG | | | |
| n | 70 | 71 | - |
| GMT in BAU/ml | 1822 | 2381 | 0.765 |
| (95% CI) | (1451-2289) | (1937-2927) | (0.593-0.987) |
| Neutralizing antibodies (Wuhan) | | | |
| n | 70 | 71 | - |
| $NT_{50}$ | 216 | 279 | 0.776 |
| (95% CI) | (170-276) | (224-347) | (0.592-1.017) |
| Neutralizing antibodies (Delta) | | | |
| n | 66 | 70 | - |
| $NT_{50}$ | 50 | 52 | 0.965 |
| (95% CI) | (40-62) | (43-63) | (0.758-1.227) |

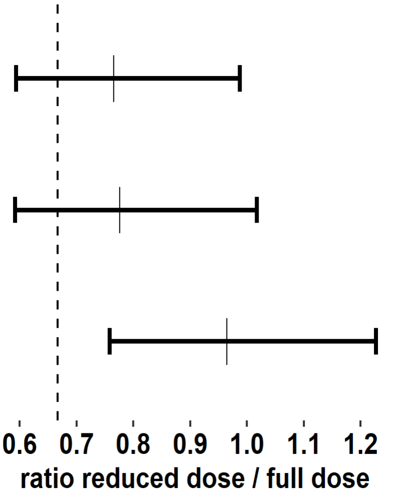


Data are geometric mean titres (95% CI) at day 28 post second dose. GMRs (95% CI) were adjusted with a linear mixed-effect model including gender, age and SARS-CoV-2 anti-RBD IgG titre at baseline as fixed variables and location as random variable. In the figure, the dashed line indicates the WHO recommended non-inferiority margin of 0.67. GMT=geometric mean titre. GMR=geometric mean ratio. BAU=Binding antibody units. NT50=50% neutralizing antibody titre.

**S2 Table. Humoral and cellular responses by study arm in the intention-to-treat cohort at the different time points.**

| | Intention-to-treat | | |
|---|---|---|---|
| | 20 µg | 30 µg | p-value |
| **SARS-CoV-2 anti-RBD IgG** | | | |
| **Day 0** | | | |
| n | 70 | 71 | |
| Concentration (BAU/ml) | 10.9 | 9.9 | p=0.310 |
| | (9.1-12.9) | (8.3-11.8) | |
| > 5 BAU/ml | 12 | 10 | p=0.649 |
| | (17%, 9-28) | (14%, 7-24) | |
| **Day 21** | | | |
| n | 70 | 71 | |
| Concentration (BAU/ml) | 173 | 296 | p=0.001 |
| | (125-240) | (218-400) | |
| > 5 BAU/ml | 69 | 71 | p=0.497 |
| | (99% 92-100) | (100%, 95-100) | |
| **Day 49** | | | |
| n | 70 | 71 | |
| Concentration (BAU/ml) | 1822 | 2381 | p=0.040 |
| | (1451-2289) | (1937-2927) | |
| > 5 BAU/ml | 70 | 71 | p=1.000 |
| | (100%, 95-100) | (100%, 95-100) | |
| **Month 6** | | | |
| n | 59 | 67 | |
| Concentration (BAU/ml) | 190 | 272 | p=0.019 |
| | (143-254) | (212-349) | |
| > 5 BAU/ml | 59 | 67 | p=1.000 |
| | (100%, 94-100) | (100%, 95-100) | |
| **Neutralizing antibodies** | | | |
| **Wuhan - Day 21** | | | |
| N | 70 | 71 | |
| $NT_{50}$ | 45 | 47 | p=0.677 |
| | (36-56) | (39-58) | |
| ➢ 50 | 17 | 16 | p=0.844 |
| | (24%, 15-36) | (23%, 13-34) | |
| **Wuhan - Day 49** | | | |
| N | 70 | 71 | |
| $NT_{50}$ | 216 | 279 | p=0.066 |
| | (170-276) | (224-347) | |
| > 50 | 66 | 70 | p=0.209 |
| | (94%, 86-98) | (99%, 92-100) | |
| **Delta - Day 49** | | | |
| n | 66 | 70 | - |
| $NT_{50}$ | 50 | 52 | p=0.767 |
| | (40-62) | (43-63) | |
| > 50 | 31 | 31 | p=0.863 |
| | (47%, 35-60) | (44%, 32-57) | |
| **Omicron - Day 49** | | | |
| n | 18 | 17 | |
| $NT_{50}$ | 37 | 40 | p=0.612 |
| | (28-48) | (30-54) | |
| > 50 | 8 | 5 | p=0.489 |
| | (44%, 22-69) | (29%, 10-56) | |

**S2 Table (continued). Humoral and cellular responses by study in the intention-to-treat cohort at the different time points .**

| | Intention-to-treat | | |
|---|---|---|---|
| | 20 µg | 30 µg | p-value |
| **IFN-γ producing cells per million PBMCs (ELISpot)** | | | |
| **SARS-CoV-2 S1-specific - Day 49** | | | |
| n | 19 | 26 | |
| Mean number of cells/million | 72.7 (35.8-147.8) | 116.5 (67.3-201.6) | p=0.219 |
| > LOD | 13 (68%, 43-87) | 21 (81%, 61-93) | p=0.818 |
| **SARS-CoV-2 S2-specific - Day 49** | | | |
| n | 19 | 26 | |
| Mean number of cells/million | 96.4 (39.8-233.1) | 127.4 (58.1-279.1) | p=0.494 |
| > LOD | 12 (63%, 38-84) | 18 (69%, 48-86) | p=1.000 |
| **SARS-CoV-2 specific T-cells (Flow cytometry)** | | | |
| **CD4+ (S1) – Day 49** | | | |
| n | 19 | 26 | |
| % | 0.086 (0.050-0.147) | 0.105 (0.063-0.177) | p=0.367 |
| > 0.0001% | 19 (100%, 82-100) | 26 (100%, 87-100) | p=1.000 |
| **CD8+ (S1) - Day 49** | | | |
| N | 19 | 26 | |
| % | 0.010 (0.001-0.113) | 0.011 (0.001-0.116) | p=0.920 |
| > 0.0001% | 14 (74%, 49-91) | 16 (62%, 41-80) | p=0.813 |
| **CD4+ (S2) - Day 49** | | | |
| N | 19 | 26 | |
| % | 0.092 (0.051-0.165) | 0.133 (0.076-0.235) | p=0.140 |
| > 0.0001% | 19 (100%, 82-100) | 26 (100%, 87-100) | p=1.000 |
| **CD8+ (S2) - Day 49** | | | |
| N | 19 | 26 | |
| % | 0.035 (0.004-0.339) | 0.038 (0.004-0.345) | p=0.931 |
| > 0.0001% | 13 (68%, 43-87) | 18 (69%, 48-86) | p=1.000 |

Data are geometric means (95% CI) for continuous variables, and n (%, 95% CI) for binary values. The LLOQ for the SARS-CoV-2 anti-RBD IgG titre was 5 BAU/mL, NT50=50 for the live virus neutralization assay, 54 and 66 cells/million PBMCs for S1 and S2, respectively, for ELIspot, and 0.0001% for flow cytometry. The mean in IFN-γ ELISpot was obtained from three replicate values. For continuous variables, p-values are reported using a linear mixed-effect model adjusted for gender, age and baseline infection status (for day 0 data) or baseline SARS-CoV-2 anti-RBD IgG titre (for day 21/49 and month 6 data) as fixed variables and location as random variable. Fisher's exact test was used to report p-values for binary variables. BAU=Binding antibody units. NT50=50% neutralizing antibody titre.

A

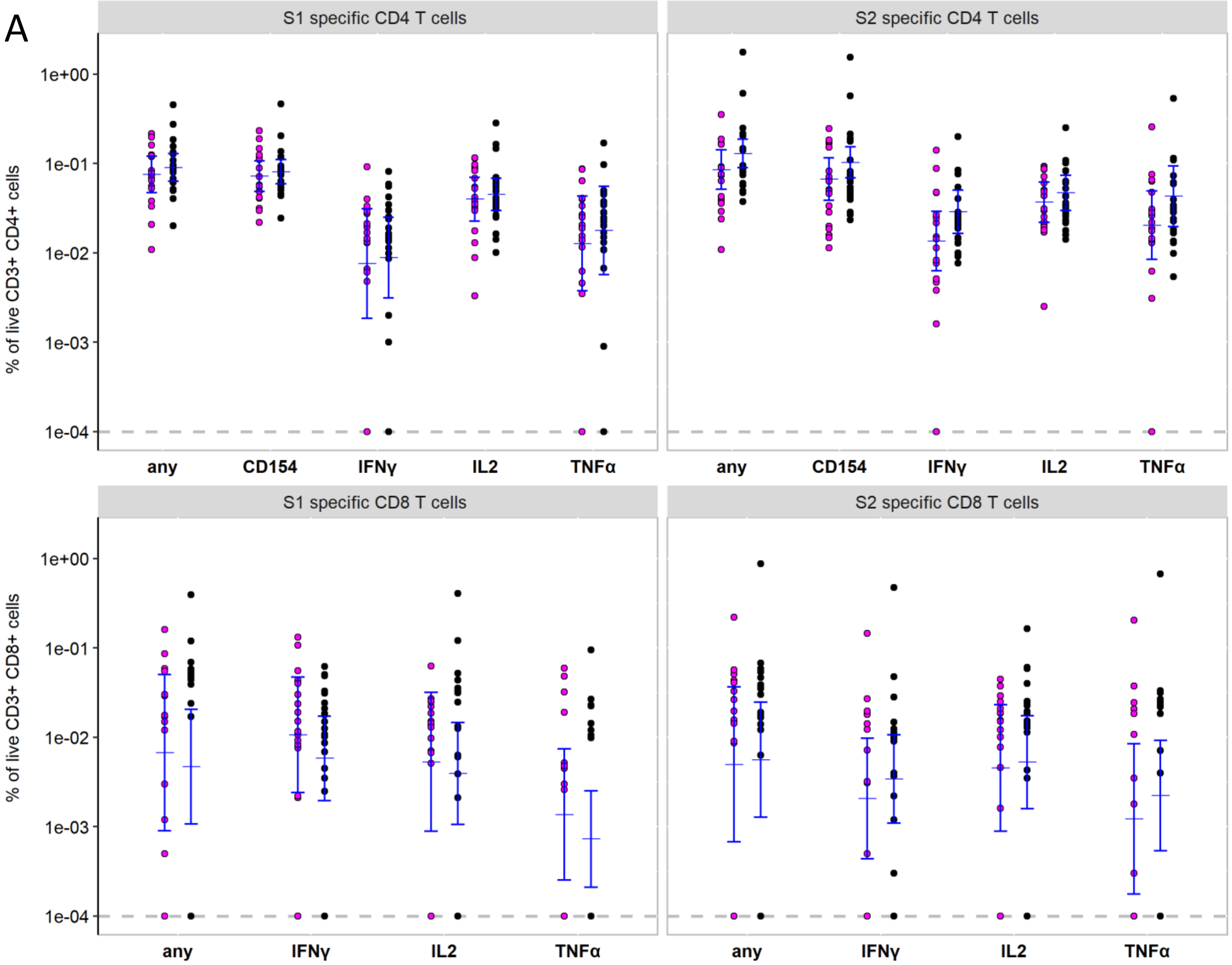


B

| | 20 µg | 30 µg | p-value | | 20 µg | 30 µg | p-value |
|---|---|---|---|---|---|---|---|
| **CD4+ (S1) - Day 49** | | | | **CD4+ (S2) - Day 49** | | | |
| n | 18 | 24 | | n | 18 | 24 | |
| % | 0.075 (0.047-0.120) | 0.090 (0.063-0.129) | p=0.447 | % | 0.085 (0.052-0.142) | 0.130 (0.089-0.188) | p=0.116 |
| > 0.0001% | 18 (100%, 81-100) | 24 (100%, 86-100) | p=1.000 | > 0.0001% | 18 (100%, 81-100) | 24 (100%, 86-100) | p=1.000 |
| **CD8+ (S1) - Day 49** | | | | **CD8+ (S2) - Day 49** | | | |
| n | 18 | 24 | | n | 18 | 24 | |
| % | 0.007 (0.001-0.050) | 0.005 (0.001-0.021) | p=0.732 | % | 0.005 (0.001-0.037) | 0.006 0.001-0.025) | p=0.907 |
| > 0.0001% | 14 (78%, 52-94) | 14 (58%, 37-78) | 0.628 | > 0.0001% | 12 (67%, 41-87) | 16 (67%, 45-84) | p=1.000 |

**S2 Fig. Flow cytometry cellular data in the per-protocol cohort at day 49 (28 days after second dose).** SARS-CoV-2 spike protein subunit 1 and 2 (S1 and S2) specific T cell frequencies were measured in 42 infection naïve participants. A. Percentage of CD4+ and CD8+ T cells expressing CD154 (only in CD4), IFN-γ, IL-2, and TNF-α are depicted. Any: percentage of cells positive for at least on the activation markers. A circle represents one test subject; the GMTs (95% CI) are shown in blue. LLOQ was fixed to 0.0001 and represented by a grey dashed line. B. Percentages are given for CD4+ and CD8+ T cells stimulated with S1 Wuhan or S2 Wuhan expressing at least one of the activation markers (corresponding to "any"). For continuous variables, p-values are reported using a linear mixed-effect model adjusted for gender, age and baseline SARS-CoV-2 anti-RBD IgG titre as fixed variables and location as random variable. Fisher's exact test was used to report p-values for binary variables.

#### S3 Table. Breakthrough infections

| # | Baseline status | Time after 1st dose (months) | Symptom duration (days) | Anti-RBD IgG (BAU/mL) Day 49 | NT50 (Wuhan) Day 49 | NT50 (Delta) Day 49 | Anti-RBD IgG (BAU/mL) Month 6 |
|---|---|---|---|---|---|---|---|
| **20µg study arm** | | | | | | | |
| 1 | Naïve | 5.2 | 14 | 3886 | 283 | 50 | **7601** |
| 2 | Naïve | 6.3 | 7 | 525 | 25 | N/A | 23 |
| 3 | Naïve | 7.1 | 12 | 603 | 66 | 25 | 58 |
| 4 | Naïve | 6.2 | 14 | 2816 | 429 | 66 | **1732** |
| 5 | Naïve | - | 0 | 616 | 75 | 25 | **2787** |
| 6 | Naïve | 6.3 | 6 | 1516 | 151 | 25 | N/A |
| 7 | Naïve | 7.1 | 3 | 2097 | 244 | 25 | 61 |
| 8 | Naïve | 5.4 | 8 | 797 | 126 | 25 | **1264** |
| 9 | Naïve | 6.9 | 5 | 694 | 82 | 25 | 131 |
| 10 | Naïve | 6.2 | 8 | 4324 | 504 | 79 | N/A |
| Average* (BTI) | | 6.3 (6.2-6.9) | 8.0 (6.0-12.0) | 1438 (763-2712) | 150 (71-316) | 36 (23-54) | 57 (18-178) |
| Average* (non-BTI) | | / | / | 1830 (1448-2311) | 163 (129-205) | 40 (33-48) | 166 (126-218) |
| **30µg study arm** | | | | | | | |
| 11 | Naïve | 6.9 | 5 | 9722 | 566 | 107 | 769 |
| 12 | Naïve | 7.3 | 3 | 3160 | 320 | 100 | 214 |
| 13 | Naïve | 6.5 | 12 | 2744 | 184 | 25 | 422 |
| 14 | Naïve | 6.8 | 10 | 1578 | 174 | 25 | 224 |
| 15 | Naïve | 6.2 | 12 | 3609 | 350 | 67 | **434** |
| 16 | Naïve | 7.2 | N/A | 1455 | 123 | 25 | 139 |
| 17 | Naïve | 6.3 | 2 | 1554 | 141 | 25 | 149 |
| 18 | Naïve | 6.3 | 7 | 2496 | 200 | 25 | 197 |
| Average* (BTI) | | 6.7 (6.3-7.0) | 7.0 (4.0-11.0) | 2686 (1593-4528) | 227 (147-350) | 40 (23-71) | 251 (143-443) |
| Average* (non-BTI) | | / | / | 2370 (1941-2894) | 213 (176-257) | 40 (34-48) | 241 (194-298) |

Subjects with a breakthrough infection (BTI) after day 49 per study arm. All BTIs were confirmed with a positive molecular test, except for subject 5 who remained asymptomatic and had a strongly elevated anti-RBD IgG titre at month 6, revealing a BTI. Binding (anti-RBD IgG) antibody titres are given at day 49 and six months post first dose. Neutralizing (NT50) antibody titres are given at day 49 (28 days post second dose). Titres in bold red indicate a breakthrough infection between vaccination and month 6 and are excluded from GMT calculation (date of infection was considered as minimum two days before the date of the positive PCR). *Indicates the median (IQR) for 'Time after 1st dose' and 'Symptom duration', and geometric mean titre (95% CI) for 'Anti-RBD IgG' and 'NT50'. BTI=breakthrough infection. BAU=Binding antibody units.

**S4 Table. Local and systemic adverse events in intention-to-treat and naïve only cohorts.**

| | **Intention-to-treat** | | | **Naive** | | |
|---|---|---|---|---|---|---|
| | **20 µg** | **30 µg** | **p-value** | **20 µg** | **30 µg** | **p-value** |
| Participants, n | 70 | 71 | | 60 | 64 | |
| Total number of local adverse events | | | | | | |
| mild | 47 | 47 | 0.95* | 41 | 42 | 0.97* |
| moderate | 46 | 44 | | 39 | 41 | |
| severe | 5 | 6 | | 5 | 4 | |
| Total number of systemic adverse events | | | | | | |
| mild | 38 | 42 | 0.63* | 29 | 39 | 0.23* |
| moderate | 87 | 76 | | 74 | 68 | |
| severe | 44 | 37 | | 43 | 33 | |
| **Local adverse events, dose 1** | | | | | | |
| none | 27 (39%) | 29 (41%) | 0.86 | 23 (38%) | 26 (41%) | 0.86 |
| pain | | | | | | |
| mild | 23 (33%) | 24 (34%) | 1* | 19 (32%) | 22 (34%) | 0.90* |
| moderate | 16 (23%) | 15 (21%) | | 14 (23%) | 14 (22%) | |
| severe | 1 (1%) | 2 (3%) | | 1 (2%) | 1 (2%) | |
| total | 40 (57%) | 41 (59%) | 1 | 34 (57%) | 37 (58%) | 1 |
| redness | | | | | | |
| mild | 2 (3%) | 1 (1%) | 1* | 2 (3%) | 1 (2%) | 1* |
| moderate | 1 (1%) | 0 | | 1 (2%) | 0 | |
| total | 3 (4%) | 1 (1%) | 0.37 | 3 (5%) | 1 (2%) | 0.35 |
| swelling | | | | | | |
| mild | 4 (6%) | 3 (4%) | 1* | 3 (5%) | 2 (3%) | 1* |
| moderate | 3 (4%) | 2 (3%) | | 2 (3%) | 2 (3%) | |
| total | 7 (10%) | 5 (7%) | 0.56 | 5 (8%) | 4 (6%) | 0.74 |
| swollen glands | | | | | | |
| mild | 1 (1%) | 1 (1%) | 1 | 1 (2%) | 1 (2%) | 1 |
| **Local adverse events, dose 2** | | | | | | |
| none | 30 (42%) | 29 (41%) | 0.87 | 24 (40%) | 26 (41%) | 1 |
| pain | | | | | | |
| mild | 12 (17%) | 13 (18%) | 0.94* | 12 (20%) | 12 (19%) | 0.53* |
| moderate | 21 (30%) | 23 (32%) | | 17 (28%) | 21 (33%) | |
| severe | 3 (4%) | 2 (3%) | | 3 (5%) | 1 (2%) | |
| total | 36 (51%) | 38 (54%) | 0.87 | 32 (53%) | 34 (53%) | 1 |
| redness | | | | | | |
| mild | 1 (1%) | 2 (3%) | 1* | 1 (2%) | 2 (3%) | 1* |
| severe | 0 | 1 (1%) | | 0 | 1 (2%) | |
| total | 1 (1%) | 3 (4%) | 0.62 | 1 (2%) | 3 (5%) | 0.62 |
| swelling | | | | | | |
| mild | 3 (4%) | 3 (4%) | 1* | 2 (3%) | 2 (3%) | 1* |
| moderate | 3 (4%) | 3 (4%) | | 3 (5%) | 3 (5%) | |
| severe | 0 | 1 (1%) | | 0 | 1 (2%) | |
| total | 6 (9%) | 7 (10%) | 1 | 5 (8%) | 6 (9%) | 1 |
| swollen glands | | | | | | |
| mild | 1 (1%) | 0 | 1* | 1 (2%) | 0 | 1* |
| moderate | 2 (3%) | 1 (1%) | | 2 (3%) | 1 (2%) | |
| severe | 1 (1%) | 0 | | 1 (2%) | 0 | |
| total | 4 (6%) | 1 (1%) | 0.21 | 4 (7%) | 1 (2%) | 0.20 |

Fisher's exact test was used to report p-values. *test between the levels of the scale of the present symptoms (mild/moderate/severe).

**S4 Table (continued). Local and systemic adverse events in intention-to-treat and naïve only cohorts**.

| | Intention-to-treat | | | Naive | | |
|---|---|---|---|---|---|---|
| | **20 µg** | **30 µg** | **p-value** | **20 µg** | **30 µg** | **p-value** |
| Participants, n | 70 | 71 | | 60 | 64 | |
| **Systemic adverse events, dose 1** | | | | | | |
| none | 39 (56%) | 43 (61%) | 0.61 | 34 (57%) | 40 (63%) | 0.58 |
| chills | | | | | | |
| mild | 1 (1%) | 0 | 1* | 1 (2%) | 0 | 1* |
| moderate | 2 (3%) | 1 (1%) | | 2 (3%) | 0 | |
| total | 3 (4%) | 1 (1%) | 0.37 | 3 (5%) | 0 | 0.11 |
| diarrhea | | | | | | |
| moderate | 1 (1%) | 0 | 1* | 0 | 0 | 1* |
| severe | 1 (1%) | 0 | | 1 (2%) | 0 | |
| total | 2 (3%) | 0 | 0.24 | 1 (2%) | 0 | 0.48 |
| fatigue | | | | | | |
| mild | 3 (4%) | 4 (6%) | 0.11* | 1 (2%) | 4 (6%) | 0.16* |
| moderate | 12 (17%) | 5 (7%) | | 10 (17%) | 5 (8%) | |
| severe | 3 (4%) | 7 (10%) | | 3 (5%) | 5 (8%) | |
| total | 18 (26%) | 16 (23%) | 0.70 | 14 (23%) | 14 (22%) | 1 |
| fever | | | | | | |
| mild | 0 | 1 (1%) | 1 | 0 | 0 | 1 |
| flu-like symptom | | | | | | |
| mild | 0 | 1 (1%) | 1 | 0 | 1 (2%) | 1 |
| headache | | | | | | |
| mild | 2 (3%) | 5 (7%) | 0.49* | 2 (3%) | 4 (6%) | 0.62* |
| moderate | 7 (10%) | 7 (10%) | | 6 (10%) | 5 (8%) | |
| severe | 1 (1%) | 0 | | 1 (2%) | 0 | |
| total | 10 (14%) | 12 (17%) | 0.82 | 9 (15%) | 9 (14%) | 1 |
| heating/sweating | | | | | | |
| moderate | 1 (1%) | 0 | 1* | 1 (2%) | 0 | 1* |
| severe | 1 (1%) | 0 | | 1 (2%) | 0 | |
| total | 2 (3%) | 0 | 0.24 | 2 (3%) | 0 | 0.23 |
| joint pain | | | | | | |
| moderate | 1 (1%) | 0 | 0.50 | 1 (2%) | 0 | 0.48 |
| malaise | | | | | | |
| mild | 1 (1%) | 0 | 1* | 1 (2%) | 0 | 1* |
| moderate | 0 | 1 (1%) | | 0 | 1 (2%) | |
| total | 1 (1%) | 1 (1%) | 1 | 1 (2%) | 1 (2%) | 1 |
| muscle pain | | | | | | |
| mild | 4 (6%) | 2 (3%) | 1* | 3 (5%) | 2 (3%) | 1* |
| moderate | 6 (9%) | 4 (6%) | | 4 (7%) | 2 (3%) | |
| severe | 2 (3%) | 1 (1%) | | 2 (3%) | 0 | |
| total | 12 (17%) | 7 (10%) | 0.23 | 9 (15%) | 4 (6%) | 0.15 |
| nausea | | | | | | |
| mild | 1 (1%) | 2 (3%) | 1* | 1 (2%) | 1 (2%) | 1* |
| moderate | 2 (3%) | 1 (1%) | | 2 (3%) | 1 (2%) | |
| severe | 0 | 1 (1%) | | 0 | 1 (2%) | |
| total | 3 (4%) | 4 (6%) | 1 | 3 (5%) | 3 (5%) | 1 |

Fisher’s exact test was used to report p-values. *test between the levels of the scale of the present symptoms (mild/moderate/severe).

**S4 Table (continued). Local and systemic adverse events in intention-to-treat and naïve only cohorts**.

| | | Intention-to-treat | | | Naive | |
|---|---|---|---|---|---|---|
| | **20 µg** | **30 µg** | **p-value** | **20 µg** | **30 µg** | **p-value** |
| Participants, n | 70 | 71 | | 60 | 64 | |
| **Systemic adverse events, dose 2** | | | | | | |
| none | 21 (30%) | 27 (38%) | 0.38 | 17 (28%) | 23 (36%) | 0.44 |
| ageusia | | | | | | |
| moderate | 1 (1%) | 0 | 0.50 | 1 (2%) | 0 | 0.48 |
| anosmia | | | | | | |
| moderate | 1 (1%) | 0 | 0.50 | 1 (2%) | 0 | 0.48 |
| chills | | | | | | |
| mild | 3 (4%) | 2 (3%) | 0.38* | 2 (3%) | 2 (3%) | 0.51* |
| moderate | 2 (3%) | 7 (10%) | | 2 (3%) | 7 (11%) | |
| severe | 2 (3%) | 2 (3%) | | 2 (3%) | 2 (3%) | |
| total | 7 (10%) | 11 (15%) | 0.45 | 6 (10%) | 11 (17%) | 0.30 |
| decreased appetite | | | | | | |
| severe | 0 | 1 (1%) | 0.50 | 0 | 1 (2%) | 1 |
| diarrhea | | | | | | |
| mild | 0 | 1 (1%) | 1* | 0 | 1 (2%) | 1* |
| moderate | 0 | 1 (1%) | | 0 | 1 (2%) | |
| severe | 0 | 1 (1%) | | 0 | 1 (2%) | |
| total | 0 | 3 (4%) | 0.24 | 0 | 3 (5%) | 0.24 |
| fatigue | | | | | | |
| mild | 6 (9%) | 6 (8%) | 0.86* | 5 (8%) | 6 (9%) | 0.65* |
| moderate | 15 (21%) | 17 (24%) | | 13 (22%) | 16 (25%) | |
| severe | 16 (23%) | 13 (18%) | | 16 (27%) | 12 (19%) | |
| total | 37 (53%) | 36 (51%) | 0.87 | 34 (57%) | 34 (53%) | 0.72 |
| fever | | | | | | |
| mild | 4 (6%) | 4 (6%) | 1* | 3 (5%) | 4 (6%) | 1* |
| moderate | 2 (3%) | 2 (3%) | | 2 (3%) | 2 (3%) | |
| total | 6 (9%) | 6 (8%) | 1 | 5 (8%) | 6 (9%) | 1 |
| flu-like symptom | | | | | | |
| mild | 1 (1%) | 0 | 1* | 1 (2%) | 0 | 1* |
| moderate | 1 (1%) | 0 | | 0 | 0 | |
| total | 2 (3%) | 0 | 0.24 | 1 (2%) | 0 | 0.48 |
| headache | | | | | | |
| mild | 6 (9%) | 4 (6%) | 0.67* | 6 (10%) | 4 (6%) | 0.61* |
| moderate | 17 (24%) | 17 (24%) | | 14 (23%) | 16 (25%) | |
| severe | 6 (9%) | 3 (4%) | | 6 (10%) | 3 (5%) | |
| total | 29 (41%) | 24 (34%) | 0.39 | 26 (43%) | 23 (36%) | 0.46 |
| heating/sweating | | | | | | |
| moderate | 0 | 2 (3%) | 0.33* | 0 | 2 (3%) | 0.33* |
| severe | 2 (3%) | 0 | | 2 (3%) | 0 | |
| total | 2 (3%) | 2 (3%) | 1 | 2 (3%) | 2 (3%) | 1 |
| insomnia | | | | | | |
| severe | 0 | 1 (1%) | 0.50 | 0 | 1 (2%) | 1 |
| joint pain | | | | | | |
| mild | 0 | 1 (1%) | 0.20* | 0 | 1 (2%) | 0.20* |
| moderate | 3 (4%) | 0 | | 3 (5%) | 0 | |
| severe | 1 (1%) | 1 (1%) | | 1 (2%) | 1 (2%) | |
| total | 4 (6%) | 2 (3%) | 0.44 | 4 (7%) | 2 (3%) | 0.43 |

Fisher's exact test was used to report p-values. *test between the levels of the scale of the present symptoms (mild/moderate/severe).

**S4 Table (continued). Local and systemic adverse events in intention-to-treat and naïve only cohorts**.

| | | Intention-to-treat | | | Naive | |
|---|---|---|---|---|---|---|
| | **20 μg** | **30 μg** | **p-value** | **20 μg** | **30 μg** | **p-value** |
| Participants, n | 70 | 71 | | 60 | 64 | |
| **General adverse events, dose 2 (continued)** | | | | | | |
| malaise | | | | | | |
| mild | 1 (1%) | 0 | 0.40* | 0 | 0 | 1* |
| moderate | 1 (1%) | 3 (4%) | | 1 (2%) | 3 (5%) | |
| severe | 1 (1%) | 0 | | 0 | 0 | |
| total | 3 (4%) | 3 (4%) | 1 | 1 (2%) | 3 (5%) | 0.62 |
| muscle pain | | | | | | |
| mild | 5 (7%) | 5 (7%) | 1* | 3 (5%) | 5 (8%) | 0.81* |
| moderate | 7 (10%) | 8 (11%) | | 7 (12%) | 7 (11%) | |
| severe | 5 (7%) | 4 (6%) | | 5 (8%) | 4 (6%) | |
| total | 17 (24%) | 17 (24%) | 1 | 15 (25%) | 16 (25%) | 1 |
| nausea | | | | | | |
| mild | 0 | 4 (6%) | 0.01* | 0 | 4 (6%) | 0.05* |
| moderate | 4 (6%) | 0 | | 3 (5%) | 0 | |
| severe | 2 (3%) | 1 (1%) | | 2 (3%) | 1 (2%) | |
| total | 6 (9%) | 5 (7%) | 0.76 | 5 (8%) | 5 (8%) | 1 |
| palpitations | | | | | | |
| moderate | 1 (1%) | 0 | 0.50 | 1 (2%) | 0 | 0.48 |
| redness | | | | | | |
| severe | 0 | 1 (1%) | 0.50 | 0 | 1 (2%) | 1 |
| swelling | | | | | | |
| severe | 1 (1%) | 0 | 0.50 | 1 (2%) | 0 | 0.48 |

Fisher's exact test was used to report p-values. *test between the levels of the scale of the present symptoms (mild/moderate/severe).

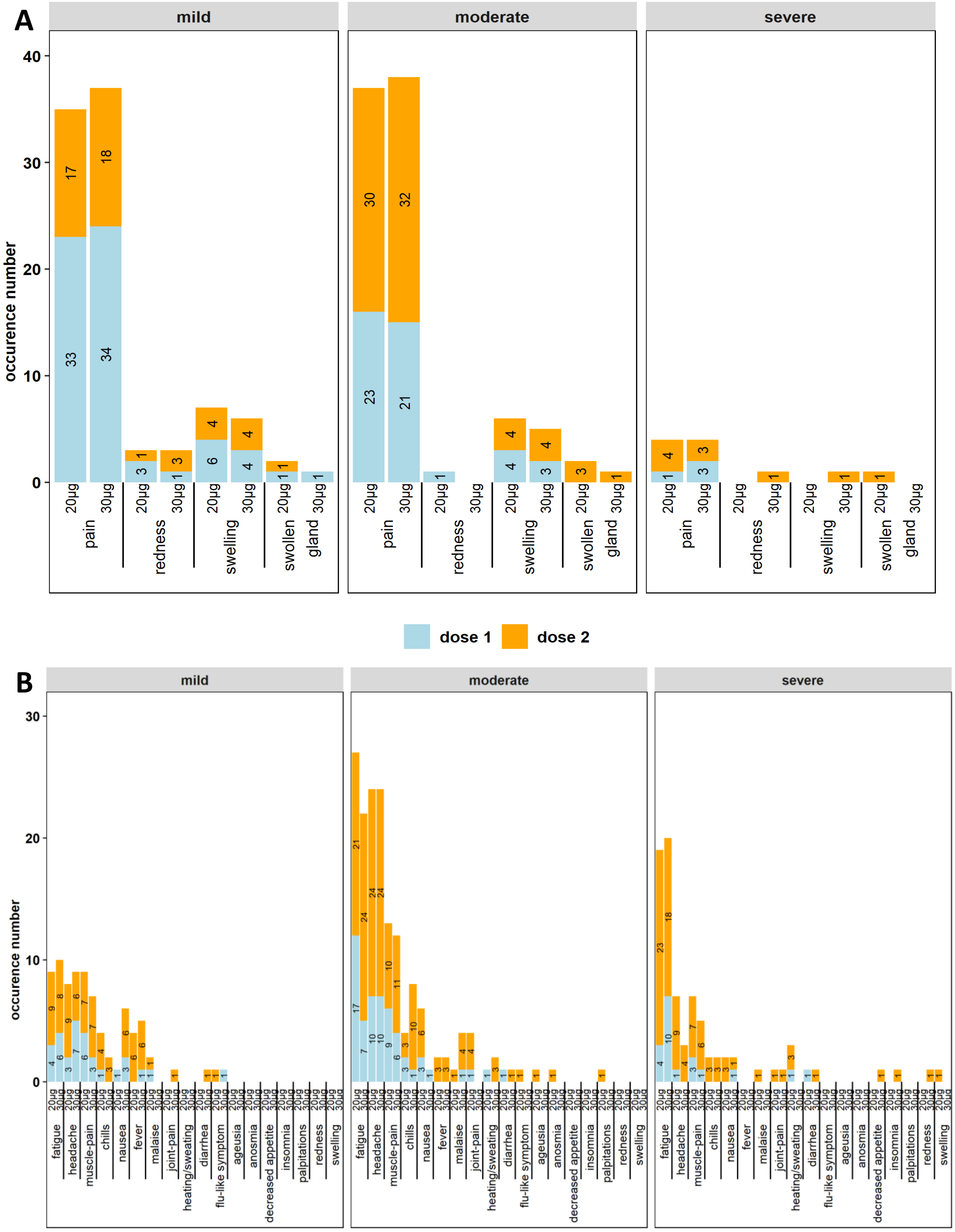


**S3 Fig. Adverse events.** Reported local (A) and systemic (B) adverse events after the first (in blue) and second (in orange) vaccine dose, according to severity (mild/moderate/severe) and by study arm (20µg and 30µg) in the intention-to-treat cohort. Occurrence number (x-axis) and percentage (numbers inside bars) per AE calculated on the cohort are given.

### S1 Appendix: Study protocol

### Safety and Immunogenicity of a Reduced Dose of the BioNTech/Pfizer BNT162b2 Vaccine in a Healthy Population (REDU-VAC)

**A randomized multicenter interventional clinical COVID-19 vaccination trial**

#### 8. Appendices

##### 8.1. APPENDIX 1: INFORMATION BROCHURE AND INFORMED CONSENT
##### 8.2. APPENDIX 2: QUESTIONNAIRE
##### 8.3. APPENDIX 3: APPROVAL BIOBANK
##### 8.4. APPENDIX 4: SUSAR FORM

### 10. Tables

**Table 1: Timetable of data and sample collection.**

| Timepoint (days) | Day of dose 1 | Day of dose 2 | 4 weeks post dose 2 | 6 months post dose 1 | 1 year post dose 1 |
|---|---|---|---|---|---|
| Informed consent | X | | | | |
| Questionnaire | X | | | X | X |
| 5 ml serum dry tube | X | X | X | X | X |
| 36 ml Heparin tubes* | X | | X | X | |

* Only for a random selection of 40 study participants, 20 per study arm.
